# Effects of rising temperature and prescribed fire on *Amblyomma Americanum* with ehrlichiosis

**DOI:** 10.1101/2022.11.07.22282052

**Authors:** Alexander Fulk, Folashade B. Agusto

**Affiliations:** Department of Ecology and Evolutionary Biology, University of Kansas, Lawrence, KS

## Abstract

Ehrlichiosis is a nationally notifiable disease in the United States and the prevalence of this disease, as with other tick-borne diseases, has been increasing since at least the year 2000. One aspect that has likely contributed to the increase in the prevalence of this disease is rising temperatures due to climate change. A promising method for control of tick populations is prescribed burning. In this study we develop a detailed mathematical model of ordinary differential equations to investigate the effect of rising temperatures on *Amblyomma americanum* populations in endemic and invasion scenarios. We used an impulsive system of ordinary differential equations to investigate the effects of prescribed burning on infectious ticks to determine if prescribed fire remains effective as temperatures increase under endemic and invasion scenarios. We found the following in the absence of prescribed burning: (1) as temperature increases, there are significant increases in the number of infectious questing nymphs and adults. (2) Ehrlichiosis becomes established in the questing nymph and adult tick populations quicker in an invasion scenario as temperature increases. In the presence of prescribed burning, we observed a reduction in the prevalence of infectious questing nymphs and adults. These results with prescribed burning hold regardless of increases in temperature, indicating that that prescribed burning remains an effective control method for *Amblyomma americanum* even in the case of a significant increase in temperature such as 2 °C and 4 °C.

## 1 Introduction

Tick-borne diseases have been increasing in incidence over the past several decades and one disease that has become of interest to researchers is human ehrlichiosis. Ehrlichiosis, formally known as human monocytic ehrlichiosis, was recognized as a human disease in the late 1980s and official reporting by the CDC on the disease began in 2000 [1, 2]. *Ehrlichia chaffeensis, Ehrlichia ewingii*, and *Ehrlichia muris eauclairensis*, are all bacterium that contribute to the diseases grouped into the term ehrlichiosis, however most cases found in the United States are caused by *Ehrlichia chaffeensis* [2]. The bacterium are transmitted to humans via their primary vector, which is *Amblyomma americanum* in the United States. This tick vector is a significant concern mainly due to its extremely aggressive and somewhat indiscriminate biting behavior in all three life stages as well as its potential for transmission of several bacterium that are known to cause disease in humans [3].

Although the number of cases of ehrlichiosis remains relatively low, they have been steadily increasing since reporting on the disease began and along with this, there has been an increase in the incidence of the disease as well [4, 5, 6]. The annual number of cases of *Ehrlichia chafeensis* ehrlichiosis in the United states when reporting began in 2000 was approximately 200 cases. As of 2019, the most recent year of publicly available annual data on the disease had over 2000 cases reported. These cases have been concentrated in several states throughout the Midwestern, Northeastern, and Southwestern United states. Some states that have consistently seen a relatively high incidence of the disease include Arkansas, Kansas, Kentucky, Missouri, North Carolina, and Tennessee [2]. This increasing incidence can be attributed to several factors including climate change, human land modification, and increased prevalence of reservoir hosts, such as white-tailed deer [7, 8]. Despite the clear threat to public health that these ticks present, their dynamics and the diseases they transmit remain relatively understudied [9, 10].

Prescribed burning is a crucial land management practice that is used to maintain certain ecosystems and reduce the risk of destructive wildfires [11, 12]. It may also be used as a possible control measure for tick populations. Overall, prescribed burning has been shown to initially reduce the prevalence of ticks in an area. The effectiveness of this control measure over longer periods of time has been debated in various field studies; however, many of the studies that have been performed to unravel the effect that prescribed burning has on ticks were flawed. Several studies focused on small plots of land that may or may not have been consistently burned prior to the study [13, 14, 15, 16, 17, 18, 19, 20]. These situations are in contrast with typical prescribed burning practices [11, 12, 21]. Often, prescribed burning is performed consistently over a long period of time on large swaths of land [21]. Gleim, et al [22] performed a study in Northwestern Florida and Southwestern Georgia that addressed the shortcomings given above and found that prescribed burning was effective at reducing the prevalence of ticks in pine and mixed pine forests. Upon further investigation, the author found that prescribed burning can even reduce the prevalence of certain tick-borne diseases [23], though this was not the case for ehrlichiosis. Further investigation is needed to determine how effective prescribed burning is at reducing the prevalence of disease in ticks and which tick-borne diseases have the potential for mitigation using this control measure.

Construction and analysis of the dynamics of mathematical models via numerical simulations allow for insights into complex situations that are difficult or impossible to simulate experimentally. Recent modeling efforts have provided further support to the effectiveness of prescribed burning as a control measure for *Ixodes scapularis* ticks [24, 25]. In addition, Wallace et. al. [26] conducted a modeling study that evaluated the effect of rising temperatures on these ticks and the primary disease that they transmit, Lyme disease. They found that rising temperature has the potential to increase transmission risk to humans due to higher tick numbers and a longer season of transmission. Thus, one of our goals is to explore the effect of rising temperature on *Amblyomma americanum* populations. Further, we aim to determine whether or not prescribed burning remains an effective control measure as temperature increases and if prescribed burning is able to reduce the percentage of infectious ticks.

An outline of the paper is as follows. Section 2 covers the formulation of the mathematical model, the impulsive system used to simulate the effect of prescribed burning, and the calculation of the parameters used to simulate the effect of prescribed burning. In Section 3 we present the results of numerical simulations covering a range of scenarios. Discussion of the numerical simulations is covered in Section 4. Finally, conclusions and future directions are given in Section 5.

## 2 Methods and modeling

### 2.1 Model formulation

To formulate the transmission model dynamics of ehrlichiosis among ticks, we adapt some of the approaches used in earlier modeling of ticks dynamics in [26, 27]. These include a complete modeling of all intermediate stages of each life-stage of the tick along with temperature-dependent development rates for certain stages and density dependent death rates for feeding ticks. We made a simplifying assumption that the number of susceptible and infectious small and large hosts is constant. We now describe each aspect in detail.

First, the number of eggs are determined, in part, by the number of egg-laying adults. The fecundity of egg-laying adults is reduced in a density-dependent manner by the factor 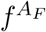, with a female having the capacity to lay a maximum of *p* = 6000 eggs if 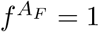. The function is given as:

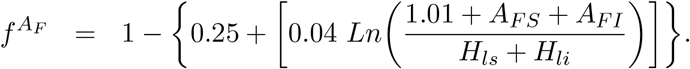

The temperature-dependent egg developmental rate is denoted as *d*_*e*_(*T*) and a they die at a fixed daily, per-capita mortality rate, *μ*_*e*_. Transovarial transmission of ehrlichiosis has long been assumed to be unlikely, if not impossible [28], and a more recent study seems to support this assumption [29], so we do not incorporate it into our model. The equation for the eggs (*E*) is given by:

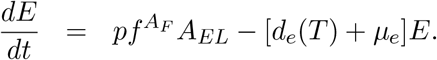

The hardening larvae population (*L*_*H*_) increases depending on the temperature-dependent egg developmental rate, *d*_*e*_(*T*). The population decreases via progression by a fixed rate *d*_*hl*_ as well as via death by the fixed rate *μ*_*hl*_.

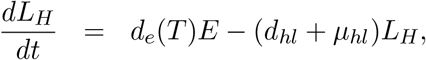

There are several components describing how the questing larvae population changes through time. First, questing larvae have a certain probability for finding a host, given by:

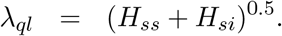

This probability depends on the total number of susceptible and infectious small hosts (*H*_*ss*_ and *H*_*si*_, respectively) and is thus fixed. We also considered the average effect that both temperature and day-length have on questing tick activity via the fixed rates *θ*_*l*_ and *δ*_*l*_, respectively. Finally, questing larvae die at the daily rate *μ*_*ql*_. The equation for the questing larvae (*L*_*Q*_) population is given by:

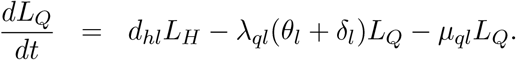

Once questing larvae find a host to feed on, they move onto the susceptible feeding larvae stage. This is the first stage where infection of a larval tick is possible if they are feeding on an infectious host, which occurs at the rate defined by the force of infection for larvae as:

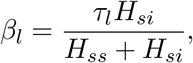

where *τ*_*l*_ is the probability of infection, *H*_*si*_ is the number of small infectious hosts, and *H*_*ss*_ is the number of small susceptible hosts. Feeding larvae also die in a density dependent manner described as:

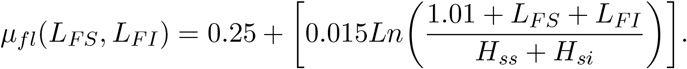

Susceptible feeding larvae (*L*_*F S*_) progress to infectious feeding larvae (*L*_*F I*_), progress to susceptible engorged larvae, or die in a density dependent manner given above as shown in the following equations:

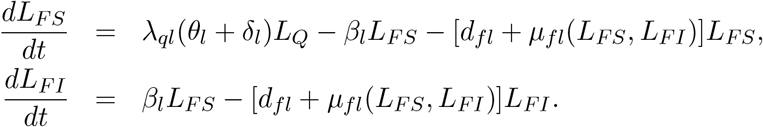

Once engorged, regardless of whether or not larvae are susceptible or infectious (*L*_*ES*_ and *L*_*EI*_, respectively), they proceed to molt into nymphs in a temperature-dependent manner, designated by *d*_*el*_(*T*), or die at the fixed rate *μ*_*el*_ as shown in the following equation:

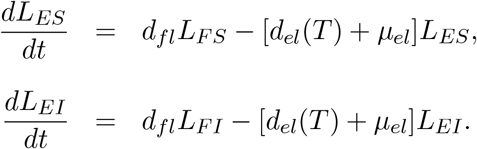

From here, a nymph will harden before questing to look for a host to feed on. Note that once a larvae (or nymph or adult) becomes infectious, it remains so for the rest of its life, as such, infected compartments for hardening nymphs are incorporated. Susceptible and infected hardening nymphs (*N*_*HS*_ and *NHI*, respectively) progress into susceptible and infectious questing nymphs at the fixed rate *d*_*hn*_ or die at the rate *μ*_*hn*_. The equations for the hardening nymphs are:

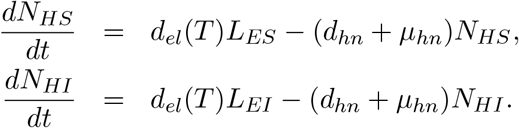

Once in the questing nymph stage, *Amblyomma americanum* ticks either die at the rate *μ*_*qn*_ or begin feeding on susceptible or infectious small or large hosts. The nymph life-stage is the least discriminant when it comes to host preference and as such, we incorporate the potential for nymphs to feed on both small and large hosts. The host finding probability for questing nymphs is thus given as:

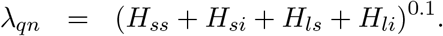

Questing activity of nymphs is affected by both temperature and day-length and we incorporate the average effects of each via the fixed parameters *θ*_*n*_ and *δ*_*n*_, respectively. Finally, the equations for susceptible and infectious questing nymphs (*N*_*QS*_ and *N*_*QI*_, respectively) is given by:

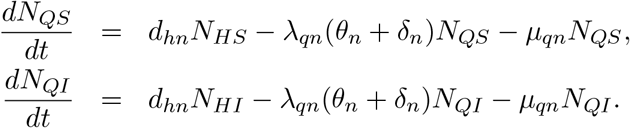

Once those questing ticks that have survived find a host, they attach and begin feeding, progressing into the feeding nymph stage. We again have at this stage that susceptible nymphs may become infected if feeding on an infectious host. The force of infection, *β*_*n*_, is defined as:

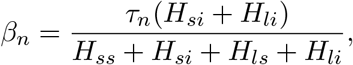

where *τ*_*n*_ is the probability that a susceptible nymph becomes infected after feeding on an infectious host. *H*_*ss*_, *H*_*si*_, *H*_*ls*_, and *H*_*li*_ are the number of susceptible and infectious small and large hosts. If susceptible feeding nymphs do not become infected, they, as well as the infectious feeding nymphs, either progress into engorged nymphs at the rate *d*_*fn*_, or die in a density-dependent manner described as:

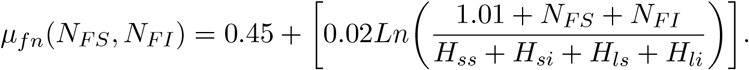

The equations describing the rates of change for susceptible and infectious feeding nymph populations (*N*_*F S*_ and *N*_*F I*_, respectively) are:

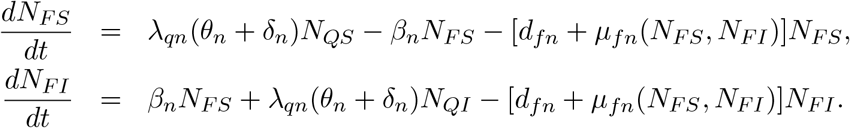

Those susceptible and infectious nymphs that survive feeding on a host then become engorged. Once engorged, they either die at a fixed rate *μ*_*en*_, or molt into hardening adult ticks based on temperature as shown in the following equations:

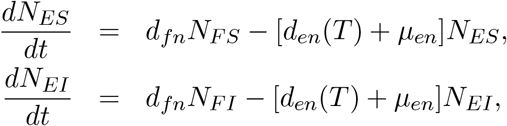

where *N*_*ES*_ denotes the susceptible engorged nymphs and *N*_*EI*_ denotes the infected engorged nymphs. Hardening adult ticks follow a similar form to hardening nymphs in that there are susceptible and infected populations of these ticks and they progress at a fixed rate, *d*_*ha*_ into susceptible or infectious questing adults. Alternatively, they die at the fixed rate *μ*_*ha*_. The two equations for hardening adults (*A*_*HS*_ and *A*_*HI*_, respectively) are:

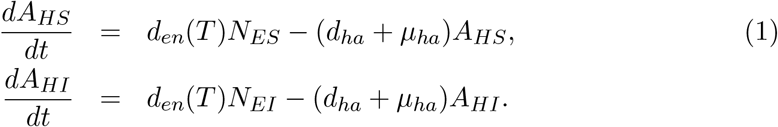

Once at the questing stage, susceptible and infectious adult ticks seek out large hosts with the following probability of success:

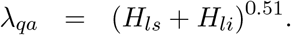

Questing activity of adult ticks is influenced by the average effects of temperature and day-length via *θ*_*a*_ and *δ*_*a*_, respectively. Susceptible and infectious questing adults may fail to find a host and die at the rate *μ*_*qa*_.The equations for adult ticks (*A*_*QS*_ and *A*_*QI*_, respectively) are guven as:

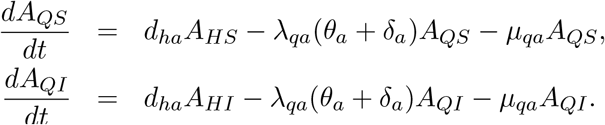

Those susceptible and infectious questing adult ticks that do find hosts progress to the feeding stage. At this point, as with the other tick life stages, we have the possibility of a tick becoming infected if feeding on an infectious host. The force of infection for feeding adult ticks is:

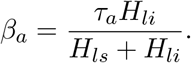

Once attached to a host, the adult ticks can take up to several days to finish feeding and progress into susceptible and infected engorged adults at the rate *d*_*fa*_. They may also die in a density dependent manner:

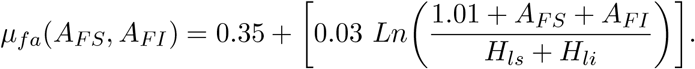

With these, we have the following equations for susceptible and infectious feeding adults, given as *A*_*F S*_ and *A*_*F I*_:

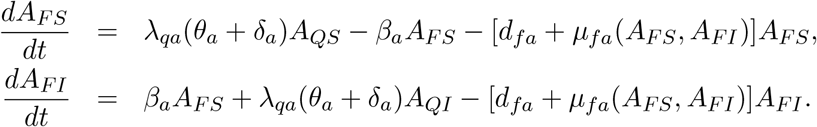

Once engorged, the female adults (*A*_*ES*_ and *A*_*EI*_) will progress into egg-laying adults at the temperature dependent rate *d*_*ea*_(*T*), or die at the rate *μ*_*ea*_ as given below:

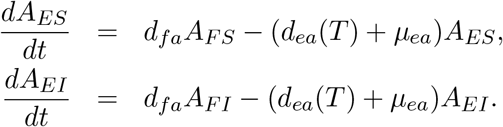

As stated previously, since transovarial transmission seems very unlikely for ehrlichiosis, we collapse egg-laying adults into a single population (*A*_*EL*_). We assume that once the eggs have been laid, all of these adults die:

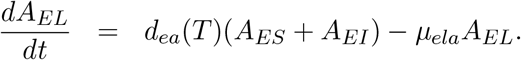

The temperature dependent development rates for eggs (*d*_*e*_(*T*)), engorged larvae (*d*_*el*_(*T*)), engorged nymphs (*d*_*en*_(*T*)), and engorged adults (*d*_*ea*_(*T*)) are given by:

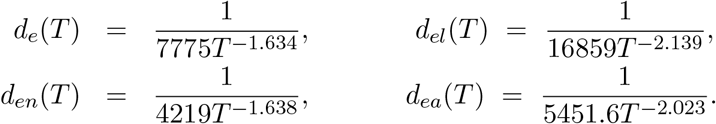

These rates were adapted from [27] and transformed into the reciprocal based on the recommendation in [30]. These rates depend on the following 5-term Fourier series function of temperature.

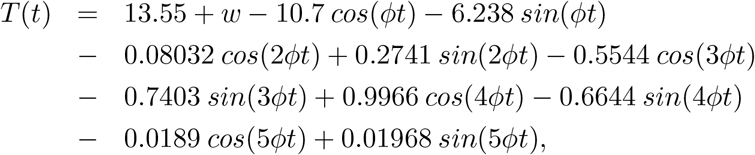

where *w* = 0, 2, 4,*° C*, and *ϕ* = 0.01826. This temperature function was created using the curve fitting toolbox in Matlab [31]. To generate the data used for calculating the temperature function, we downloaded the mean temperature data for each county in Kansas from January 1^*st*^, 2020, to December 31^*st*^, 2020 and averaged over all of those counties to get the average temperature for Kansas for that year. We removed a day before using the curve fitting toolbox since 2020 was a leap year to get a periodic function of temperature with a period equal to 365 days. The mean temperature data for each county in Kansas was downloaded from the Parameter-elevation Regressions on Independent Slopes Model (PRISM) website [32].

With these assumptions we have the following equations for the eggs and the different stages for the larvae as well as their host seeking behavior:

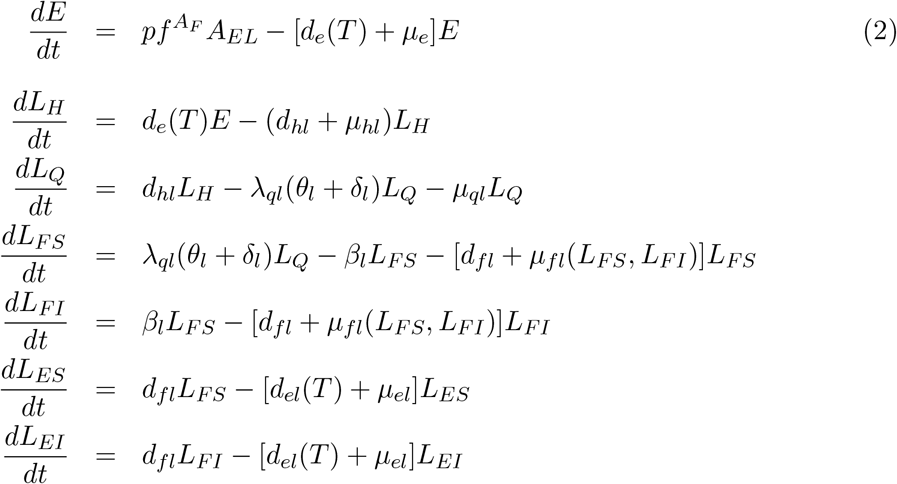

Below are the equations for the nymphs with their different life stages and host seeking behavior:

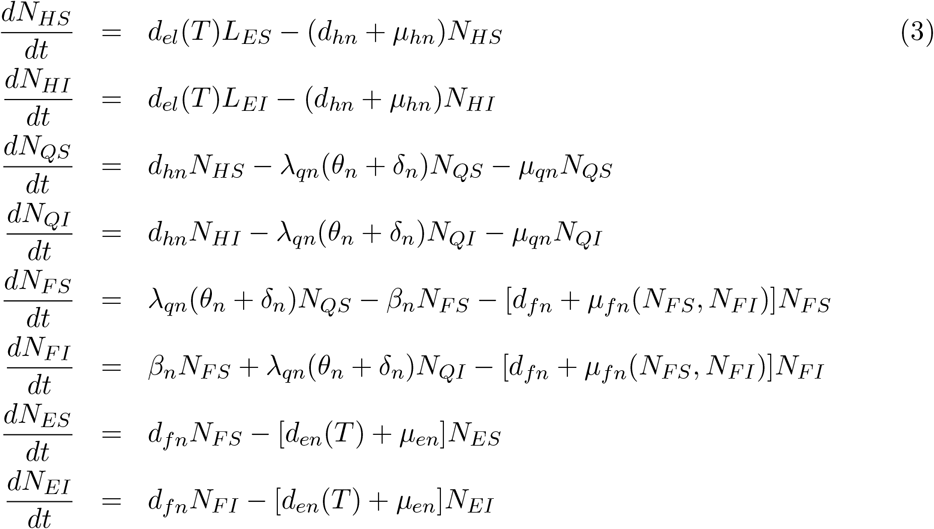

Finally, we provide the equations for the adult tick populations that are described in detail above:

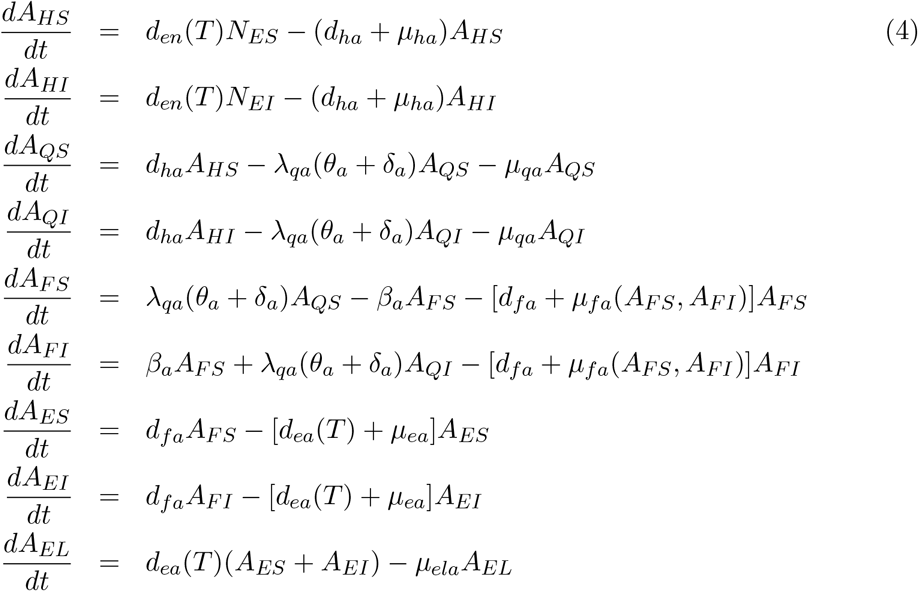

The model flow diagram is given in Figure 1 and the description of the parameter and the values used in simulations are given in Table 1. Note the arrows from the purple box in Figure 1 signifies the temperature-dependent developmental rates.

**Table 1:** Description of the variables and parameters for ehrlichiosis transmission model (2) - (4) in *Amblyomma americanum*. Parameter values and estimates are taken from [27].

| Parameter | Meaning | Value |
| --- | --- | --- |
| $p$ | Birthrate of tick eggs | 6000 |
| $\mu_e$ | Death/inviability rate of eggs | 0.008 |
| $d_{hl}$ | Progression rate of hardening larva | 1/5 |
| $\mu_{hl}$ | Death rate of hardening larva | 0.00648 |
| $\theta_l$ | Average effect of temperature on activity of questing larva | 0.002 |
| $\delta_l$ | Average effect of day-length on activity of questing larva | 0 |
| $\mu_{ql}$ | Death rate of questing larva | 0.0029 |
| $\tau_l$ | Probability of infection in larva | 0.1 |
| $d_{fl}$ | Progression rate of feeding larva | 1/5 |
| $\mu_{el}$ | Death rate of engorged larva | 0.005 |
| $d_{hn}$ | Progression rate of hardening nymphs | 1/7 |
| $\mu_{hn}$ | Death rate of hardening nymphs | 0.003 |
| $\theta_n$ | Average effect of temperature on activity of questing nymphs | 0.01 |
| $\delta_n$ | Average effect of day-length on activity of questing nymphs | 0.005 |
| $\mu_{qn}$ | Death rate of questing nymphs | 0.001 |
| $\tau_n$ | Probability of infection in feeding nymphs | 0.1 |
| $d_{fn}$ | Progression rate of feeding nymphs | 1/5 |
| $\mu_{en}$ | Death rate of engorged nymphs | 0.004 |
| $d_{ha}$ | Progression rate of hardening adults | 1/14 |
| $\mu_{ha}$ | Death rate of hardening adults | 0.0035 |
| $\theta_a$ | Average effect of temperature on activity of questing adults | 0.007 |
| $\delta_a$ | Average effect of day-length on activity of questing adults | 0.004 |
| $\mu_{qa}$ | Death rate of questing adults | 0.00015 |
| $\tau_a$ | Probability of infection in feeding adults | 0.1 |
| $d_{fa}$ | Progression rate of feeding adults | 1/12 |
| $\mu_{ea}$ | Death rate of engorged adults | 0.003 |
| $\mu_{ela}$ | Death rate of egg-laying adults | 1 |

**Figure 1:**
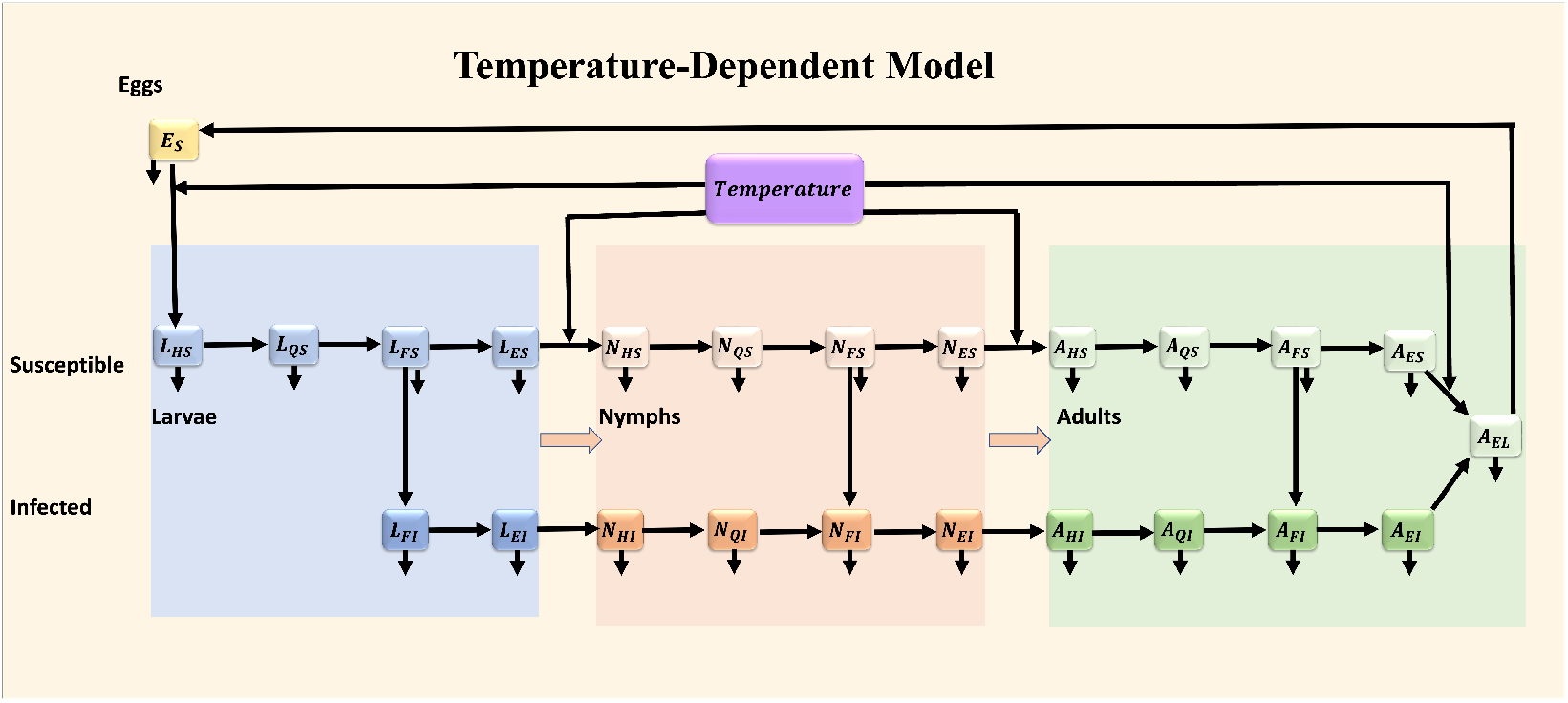
Flow diagram of ehrlichiosis transmission model (2) - (4) in *Amblyomma americanum* with temperature.

### 2.2 The impulsive system

To incorporate the effect of fire on the tick populations we use a system of impulsive differential equations. Next, we provide the impulsive component of the system starting with the egg and larvae populations. For time *t* ≠ *nT, n* = 1, 2, …, when fire is not implemented, we have the following system of equations for the eggs and larvae populations:

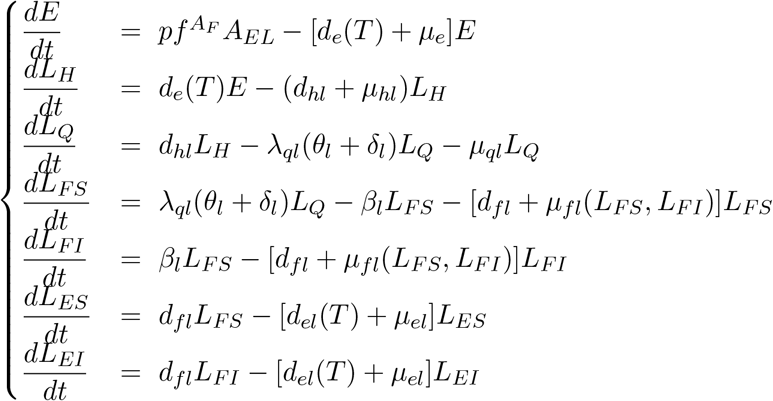

At a predetermined time *t* = *nT*, we initiate a burn that reduces each tick population by a particular proportion using the following equation:

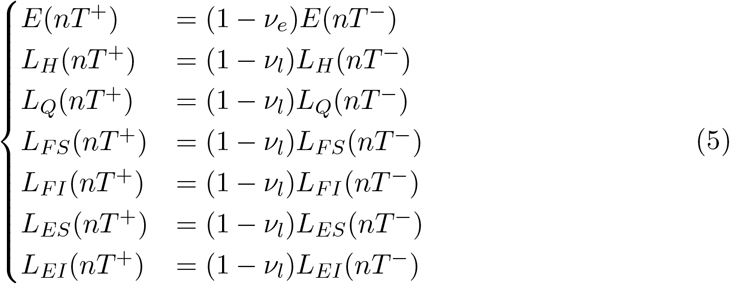

The parameters *ν*_*i*_, *i* = *e, l*, indicate a reduction in each population with values ranging from zero to one hundred percent reduction following a burn. These reduced populations are then used as the initial conditions in the model simulation until the next burn is implemented.

For the nymph populations we have the following equations in the time between burns (*t* ≠ *nT*):

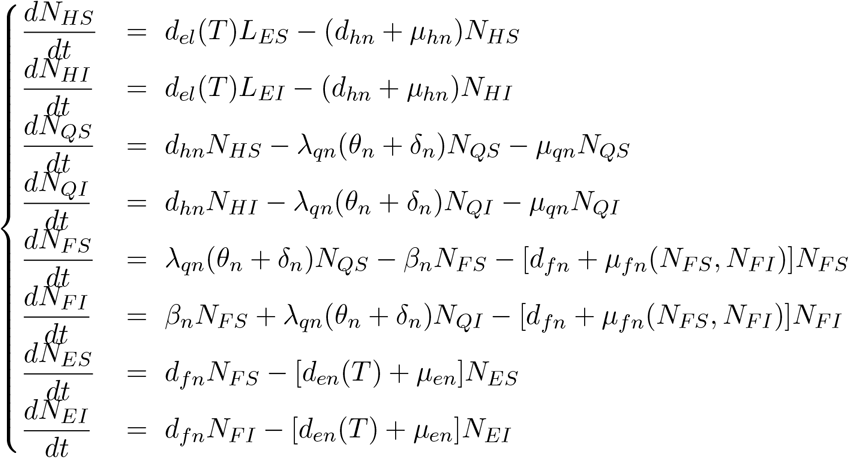

The nymph populations are reduced at the end of each interval by the proportion (1 *− ν*_*n*_) in the following manner at time *t* = *nT* :

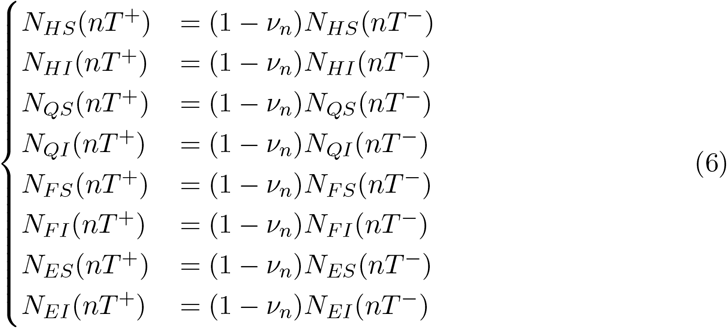

Lastly, the adult tick populations are governed by the following equations in the time between burns (*t≠ nT*):

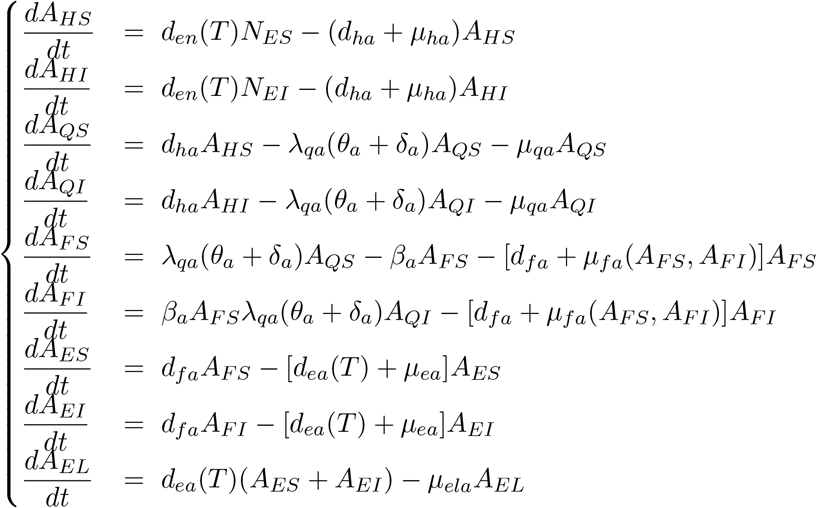

And the adult tick populations are subsequently reduced by the proportion (1 *−ν*_*a*_) when a burn occurs at time *t* = *nT* :

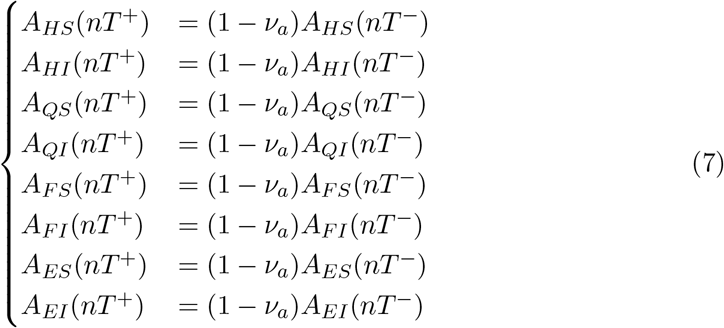

### 2.3 Estimating the proportion of ticks reduced due to a prescribed burn

Limited data exists on sampling efforts made before and after prescribed burns that might allow us to accurately determine the proportion of each tick life stage reduced following a prescribed burn. So, we used the data from Gleim et. al. [22] to calculate the proportions *ν*_*l*_, *ν*_*n*_, and *ν*_*a*_. We made the simplifying assumption that the *ν*_*e*_ parameter was zero. This in part because typical tick sampling methods are not effective at capturing tick eggs; also because tick eggs are typically laid in damp areas, such as deep in a leaf litter layer where prescribed burns may not significantly affect. For the other *ν*_*i*_, *i* = *l, n, a* parameters, we used the counts in 2011 from burned surrounded by burned sites for *Amblyomma americanum* in Gleim et. al. [22]. We divide the number of ticks in each life stage by the total number of ticks and subtract that proportion from one to get our estimate for the proportion of ticks reduced following a prescribed burn. For example, there were 21 nymphs and 319 *A. americanum* ticks collected in 2011 at the aforementioned sites, so we have *ν*_*n*_ = 1 - (21/319) *≈* 0.9341. The value for *ν*_*l*_ was 1 using this method, however we reduced this slightly to 0.99 since it is rather unlikely that all larvae are killed with a prescribed burn. This also was part of the motivation for us to explore how varying these parameters might affect the tick populations and those results are given in Section 3.2.3.

## 3 Numerical simulations

The purpose of this study is to explore the effects that rising temperature have on *A. americanum*, prevalence of the disease, and how these dynamics change when prescribed burns are implemented. Before addressing each of these goals, we perform a global sensitivity analysis in Section 3.1 to determine the impact the model parameters and prescribed burns have on both the number of infectious questing nymph and adult ticks and the prevalence of disease in the tick populations. In Section 3.2, we present several simulations used to investigate the effects of rising temperature on tick and disease dynamics in two different scenarios. Finally, in Section 3.2.3, we present the results of numerical simulations investigating the effect that a probabilistic implementation of prescribed burning has on *Amblyomma americanum*.

### 3.1 Global sensitivity analysis

The global sensitivity analysis allows us to determine which parameters have the strongest effects on the model outcome as well as the type of relationship (positive or negative) each parameter has with the model outcome. To create representative distributions for the model parameters, we use the Latin Hypercube Sampling method. For details on this method, see [33]. Essentially, we generate 1000 samples for each parameter, each generated from a particular distribution, in order to generate the Latin Hypercube Sampling matrix. This creates a multi-dimensional parameter space, from which, we pull various sets of parameters and run our model with each set to gauge the impact of each parameter using their partial rank correlation coefficients. We chose uniform distributions for most model parameters due to the lack of biological data pertaining to parameters governing tick dynamics and the diseases that they transmit. Parameters related to prescribed fire (i.e. the number of burns and the time between burns) are each taken from a Poisson distribution. The Poisson distribution for the number of burns is centered on four, while the Poisson distribution for the time between burns is centered on eighty. Any zeros generated from either of these distributions were subsequently changed to ones to avoid numerical issues implementing the burns. The outcome measure used is either the sum of the number of infectious questing nymphs and adults present at the end of the simulation or the sum of the prevalence of infectious questing nymphs and adults present at the end of the simulation.

This first global sensitivity analysis, shown in Figure 2, used the sum of the number of infectious questing nymphs and adults present at the end of the simulation as the outcome measure. In this case, we began each simulation at model equilibrium (i.e. an endemic scenario). The model dynamics had become periodically stable by *t* = 7300 days (20 years) and used the values for each population at that time as our starting point for each simulation. Significant parameters in this analysis included time between burns, number of burns, hardening nymph progression rate (*d*_*hn*_), probability of infection in nymphs (*τ*_*n*_), adult feeding rate (*d*_*fa*_), larvae feeding rate (*d*_*fl*_), egg death/inviability rate (*μ*_*e*_), and egg-laying adult death rate (*μ*_*ela*_). From those significant parameters, we find that the number of burns had by far the greatest impact on the number of infectious nymphs and adults present at the end of our simulations with a partial rank correlation coefficient less than -0.8. This negative coefficient intuitively implies that as the number of burns increases, the number of infectious nymphs and adults tends to decrease by the end of the simulation. Other significant parameters that followed this trend were *μ*_*e*_ and *μ*_*ela*_, though their impact on the model outcome was significantly less than the impact of the number of burns. This makes sense due to the fact that an increase of only one in the number of burns kills 93% of nymphs when the burn is initiated. Parameters that had a positive impact on the model outcome were time between burns, *d*_*hn*_, *τ*_*n*_, *d*_*fa*_, and *d*_*fl*_. These results imply that for nymphs, the most important means of generating new infectious questing nymphs are via progression following infection in the previous life stage as well as increasing the likelihood of becoming infected while feeding on an infectious host. These infected nymphs then move on to become infectious questing adults. However, since the probability of infection (*τ*_*a*_) is not significant for adult ticks, but the adult feeding rate (*d*_*fa*_) and egg-laying adult death rate (*μ*_*ela*_) are, it is likely that the most important role that the adult ticks play is creating the next generation of ticks to be infected rather than contributing directly to the outcome measure. Since infectious questing adults are maintained at an order of magnitude less than the infectious questing nymphs, it makes sense that it is better to repopulate those nymphs as quickly as possible following the dramatic decrease that occurs when a prescribed fire is implemented.

**Figure 2:**
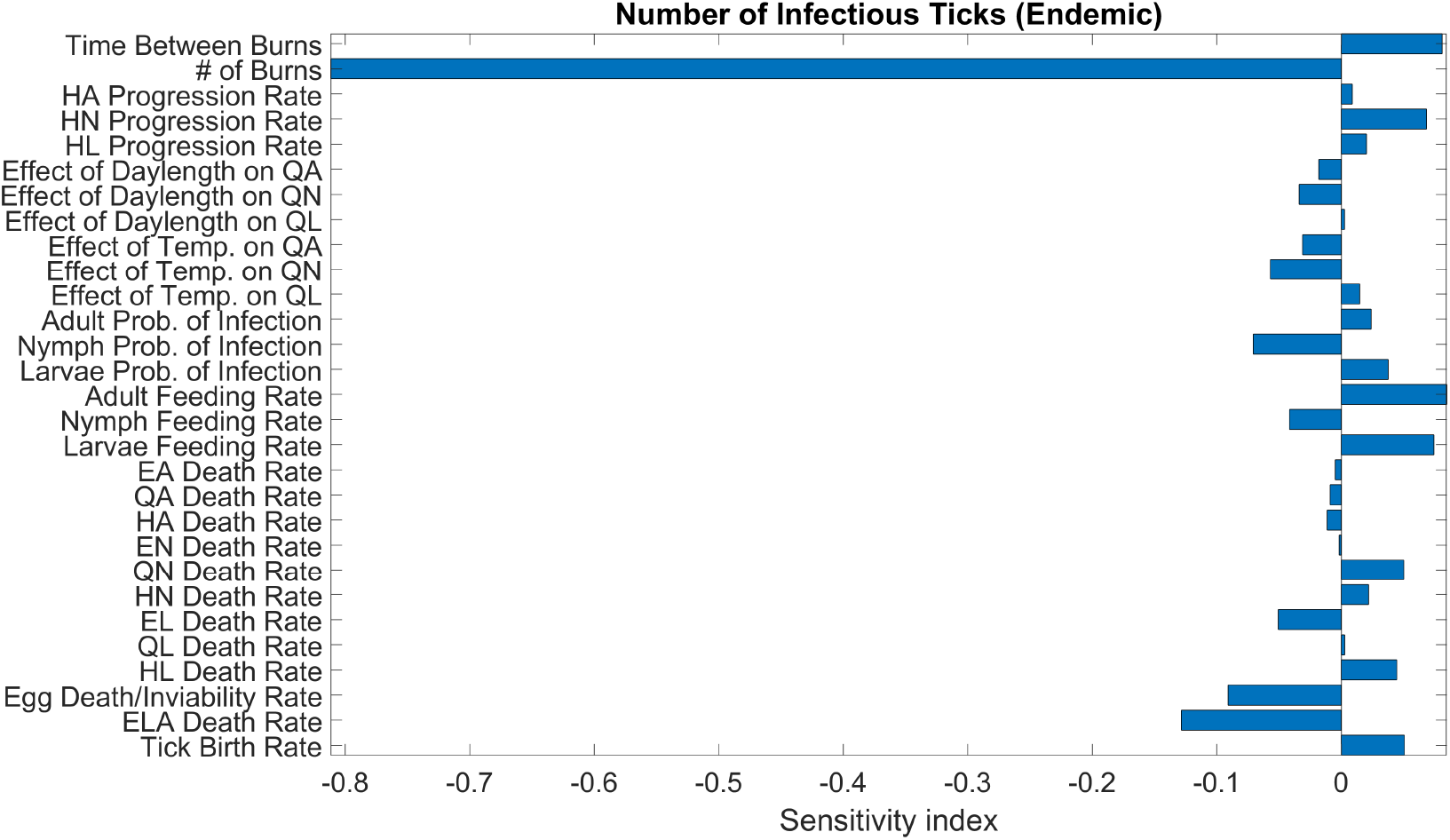
Global sensitivity analysis for ehrlichiosis transmission model (2) - (4) with temperature. In this analysis, we considered the sum of the number of infectious nymphs and adults present at the end of a simulation as our outcome measure. The simulations was carried out for an endemic scenario describe in Section 3.2.

The model outcome of a global sensitivity analysis is an important consideration as it can have dramatic effects on which parameters are significant in the analysis. Usually, the basic reproduction number, *R*_0_, is used since the goal of many modeling studies is reducing this to a value less than one which has the potential to eliminate the disease from the populations of interest. However, we assume a fixed host population and as such, we cannot calculate *R*_0_ in the usual way (i.e. using the next generation matrix method). We consider this an acceptable assumption since prescribed fire alone is unlikely to successfully reduce *R*_0_ below the necessary threshold due to the fact that it does little to affect the main reservoir host of ehrlichiosis, the white-tailed deer, other than potentially driving them from the area being burned for a short time. It is because of these reasons that we instead evaluate how the sum of the prevalence of infectious questing nymphs and adults is affected by each of the model parameters. We again use an endemic scenario as the starting place for each of our simulations. We observed in Figure 3 that the significant parameters were time between burns, number of burns, the average effect of temperature on questing nymphs (*θ*_*n*_), probability of infection in nymphs (*τ*_*n*_), probability of infection in larvae (*τ*_*l*_), and nymph feeding rate (*d*_*fn*_). From this, we again see that the number of burns has the greatest impact on the sum of the prevalence of infectious nymphs and adults, with a partial rank correlation coefficient value of -0.222. In this case, we see that the impact that this parameter has is significantly less compared to the previous outcome measure. The average effect of temperature on questing nymphs had a significant, though small, negative impact on the sum of the prevalence of infectious questing nymphs and adults. This is somewhat intuitive as the larger this value becomes, the faster infectious questing nymphs will find a host and move to the feeding nymph stage. Both *τ*_*n*_ and *d*_*fn*_ had small negative impacts on the outcome measure. The time between burns again had a significant, but small positive impact on the model outcome. The probability of infection in larvae (*τ*_*l*_) had a small positive impact on the model outcome which is unsurprising since those larvae that become infected progress to become infectious questing nymphs and adults if they survive, directly increasing the numerator of the model outcome.

**Figure 3:**
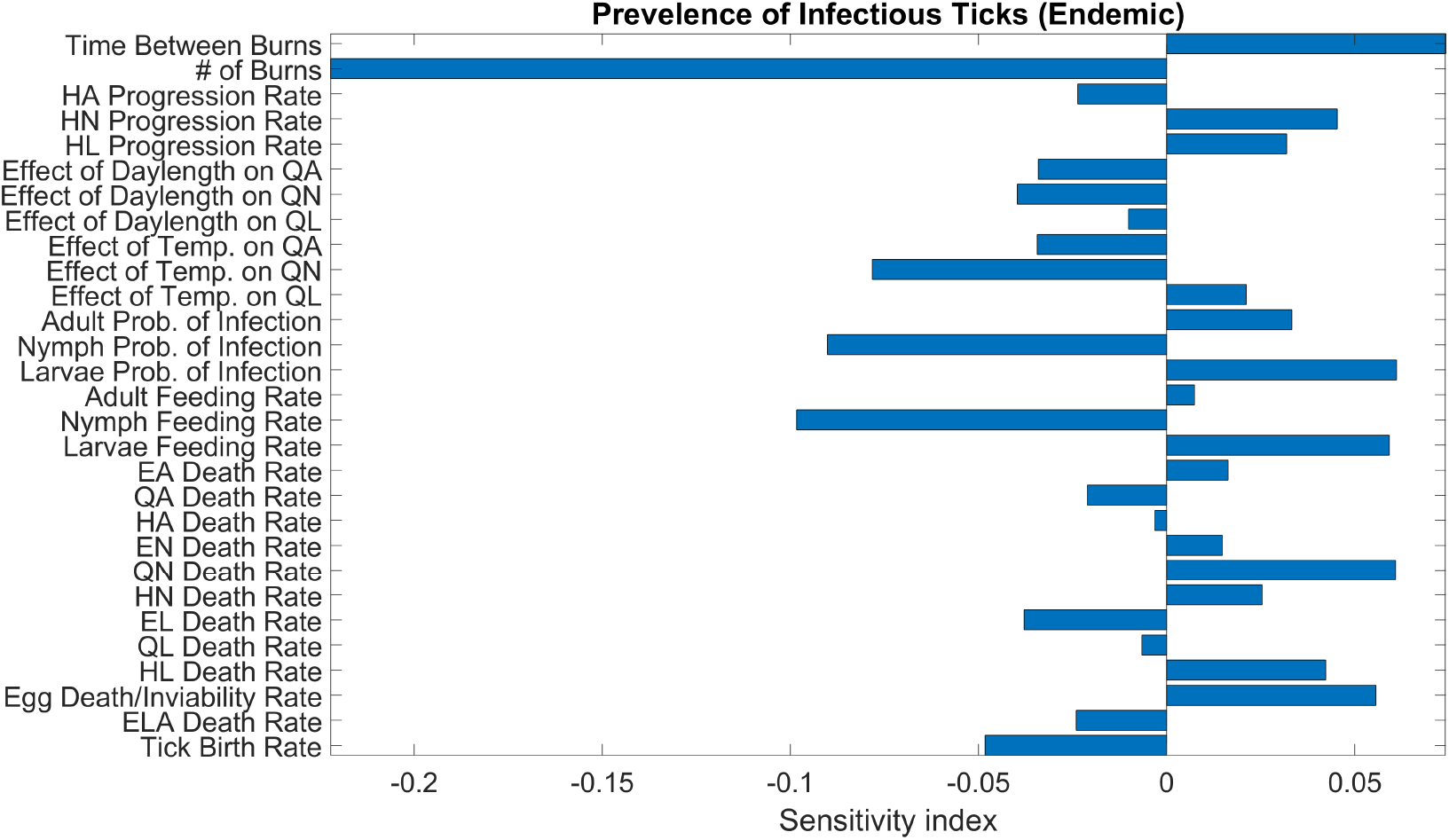
Global sensitivity analysis for ehrlichiosis transmission model (2) - (4) with temperature. In this analysis, we considered the sum of the prevalence of infectious nymphs and adults present at the end of a simulation as our outcome measure. The simulations were carried out for an endemic scenario described in Section 3.2.

Next, we evaluate the effect that model parameters have on each of these outcomes when starting at an invasion scenario in terms of the initial conditions.

Starting at the initial conditions stated for the invasion scenario described in Section 3.2.2, we ran the global sensitivity analysis for each of the outcome measures discussed previously in order to determine how the effect of each parameter changes when population and disease dynamics have not yet reached endemic dynamics. We first consider the sum of infectious questing nymphs and adults present at the end of the simulation. We found in Figure 4 that the significant parameters in this case included time between burns, number of burns, progression rate of hardening larvae (*d*_*hl*_), the average effect of temperature on questing nymphs (*θ*_*n*_), the average effect of temperature on questing larvae (*θ*_*l*_), adult feeding rate (*d*_*fa*_), larvae feeding rate (*d*_*fl*_), egg death/inviability rate (*μ*_*e*_), egg-laying adult death rate (*μ*_*ela*_), and tick birth rate (*p*). Unsurprisingly, the number of burns had the greatest negative impact on the model outcome, though the effect was not as great as it was in the endemic case. This is due to the fact that prescribed fires alone are not able to prevent the invasion of ticks and will be discussed further in Section 4. The time between burns had a much greater impact on the model outcome in this case because the growth in the number of infectious questing nymphs and adults is greater compared to the starting point near zero when compared to the endemic case. The egg death/inviability rate (*μ*_*e*_) and the egg-laying adult death rate (*μ*_*ela*_) were again significant and negative. Several parameters that increase the number of infectious questing nymphs and adults (*d*_*hl*_, *θ*_*n*_, *θ*_*l*_, *d*_*fa*_, and *d*_*fl*_) were important which implies that these rates are much more important in an invasion scenario. This makes sense as it would be easier for this tick species to become established if the environment being invaded is better suited to these ticks as would be the case if some of these parameters are increased (*d*_*hl*_, *θ*_*n*_, *θ*_*l*_). These suitable conditions also have the potential to affect the feeding rate of ticks, which generally has a positive effect on the number of infectious nymphs and adults [34].

**Figure 4:**
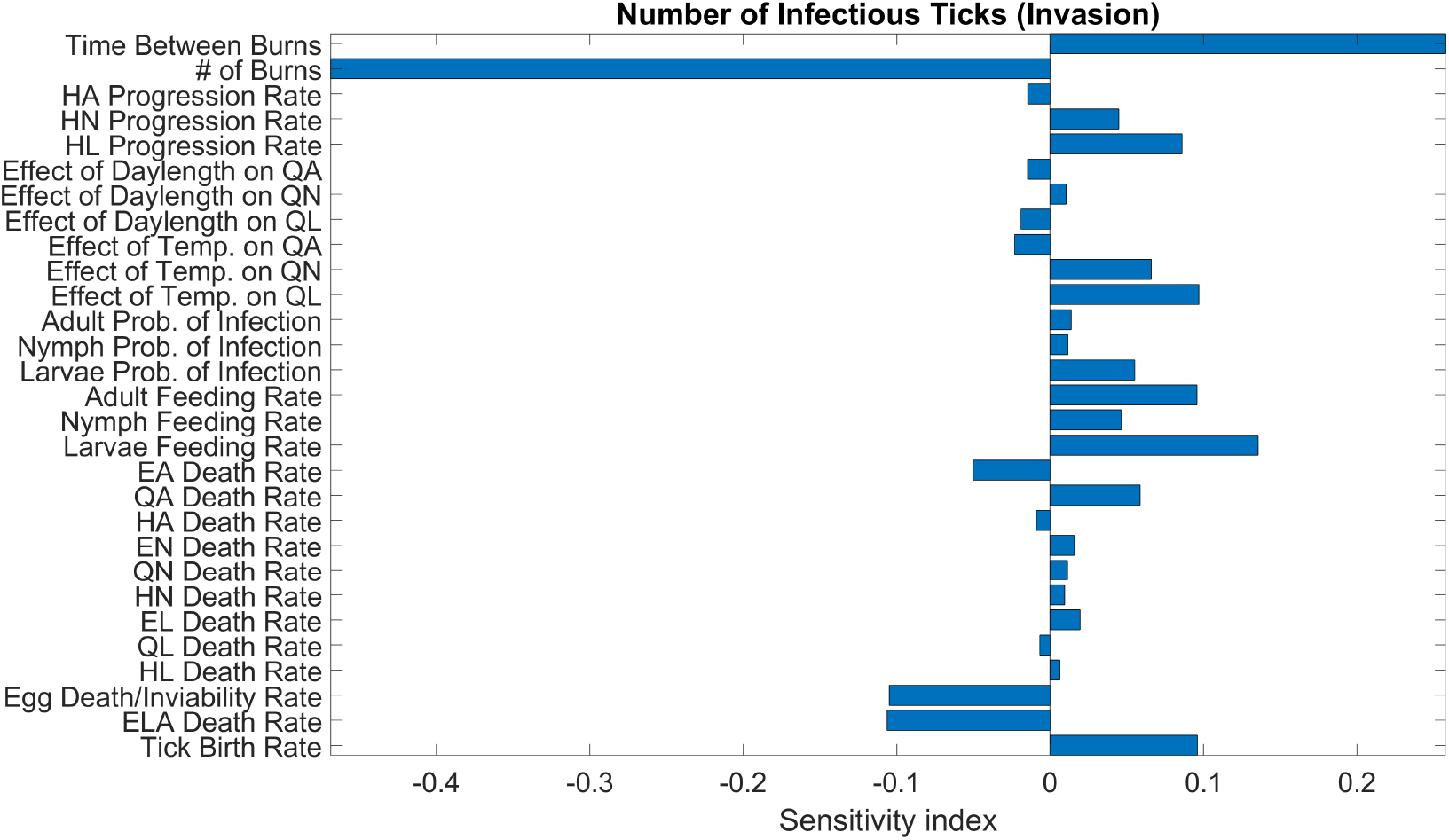
Global sensitivity analysis for ehrlichiosis transmission model (2) - (4) with temperature. In this analysis, we considered the sum of the number of infectious nymphs and adults present at the end of a simulation as our outcome measure. The simulations were carried out for an invasion scenario described in Section 3.2.

Finally, we explore how the model parameters affect the sum of the prevalence of infectious questing nymphs and adults, see Figure 5. In this case, the significant parameters were number of burns, progression rate of hardening adults (*d*_*ha*_), average effect of temperature on questing adults (*θ*_*a*_), average effect of temperature on questing larvae (*θ*_*l*_), probability of infection in larvae (*τ*_*l*_), and engorged adult death rate (*μ*_*ea*_). In contrast to the earlier global sensitivity analyses results, the parameter that had the greatest impact on the model outcome was the probability of infection in larvae (*τ*_*l*_). If this probability increases, we tend to see an increase in the prevalence of infectious questing nymphs and adults at the end of the simulation. The number of burns did have a small negative impact on the outcome, but it mattered less than the progression rate of hardening adults (*d*_*ha*_) and the engorged adult death rate (*μ*_*ea*_). Larvae and adults seem to play an important role in the disease dynamics in an invasion scenario as they have more significant parameters in this case and will be discussed further in Section 4.

**Figure 5:**
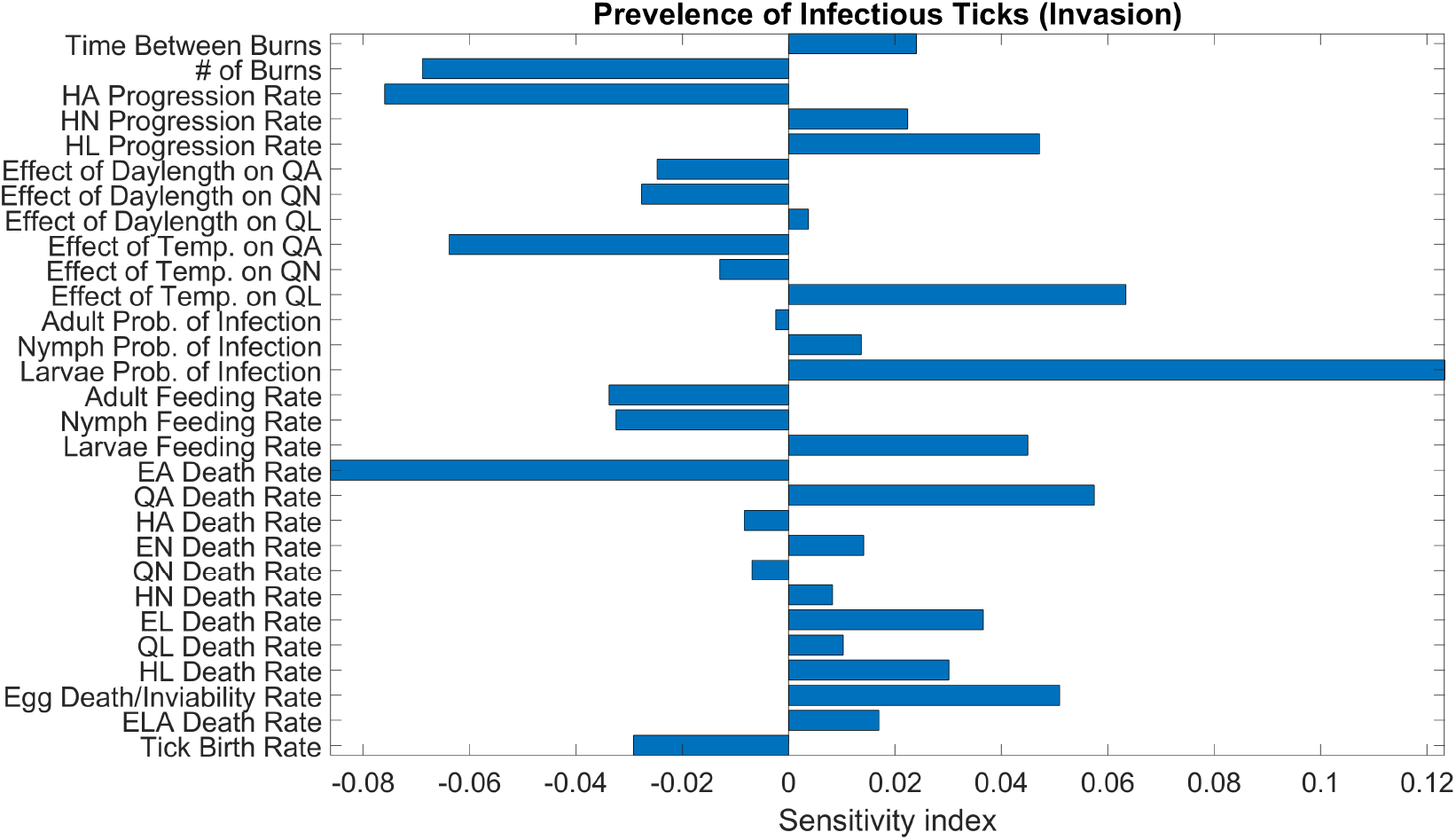
Global sensitivity analysis for ehrlichiosis transmission model (2) - (4) with temperature. In this analysis, we considered the sum of the prevalence of infectious nymphs and adults present at the end of a simulation as our outcome measure. The simulations were carried out for an invasion scenario described in Section 3.2.

### 3.2 Tick and disease dynamics in an environment with rising temperature

Next, we explore how rising temperature affects tick and disease dynamics in two different contexts that occur in many places throughout the United States, but particularly in the Midwest and throughout the Southeastern United States and also implement prescribed burns to investigate how tick populations change following a burn as temperature increases.

#### 3.2.1 Endemic dynamics

*Amblyomma americanum* are endemic throughout the North- and Southeastern United States as well as much of the Midwest. As such, we investigate the effect of rising temperature on the dynamics of questing nymph and adult tick susceptible and infectious populations at equilibrium. As stated in Section 3.1, we let the model run for 20 years, collected the values of each population at the end of the simulation and used those values as a starting point for our equilibrium simulations. The simulation presented in Figure 6 shows the model results under endemic scenario for current temperature (*w* = 0), and for increase in temperature with *w* = 2, and 4 °C.

**Figure 6:**
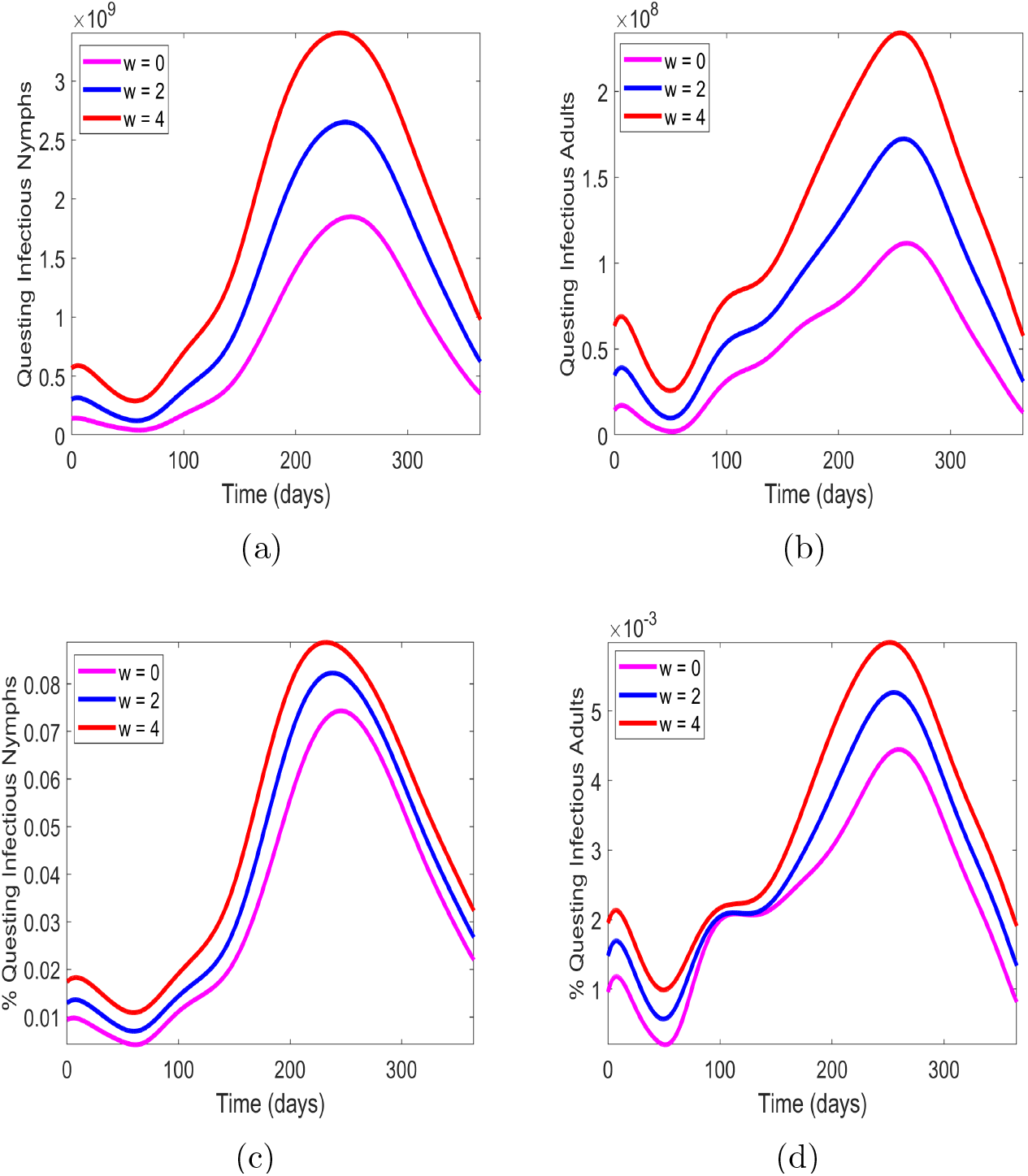
Nymph and adult tick population and disease dynamics under endemic scenario for current temperature *w* = 0 and increased temperature *w* = 2, 4 °C. (a) and (b) Questing infectious nymphs and adults; (c) and (d) Prevalence of questing nymphs and adults.

From Figure 6, we can see that as temperature increases, we have a roughly twofold increase in both the number of questing infectious nymph and adult ticks when moving from w = 0 to w = 4. We do see increases in the prevalence of ticks infected with the disease in each tick population as temperature increases, however, this still leaves a relatively low percentage in infectious questing nymphs and adults. This does not however diminish the threat of infectious ticks. Since ticks will be prevalent in much larger numbers as temperature increases and the prevalence of disease in ticks will increase slightly, the more infectious questing ticks humans are going to encounter when in areas that ticks frequent. This is also exacerbated by the fact that these increases are greatest during times when humans are likely to be outdoors in tick infested areas (July-September, 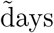 180-280). Overall, ehrlichiosis poses an increased threat to public health in areas where the disease is prevalent in ticks as temperature increases.

Next, we present the results of implementing four prescribed burns throughout the year in an endemic situation (Figure 7).

**Figure 7:**
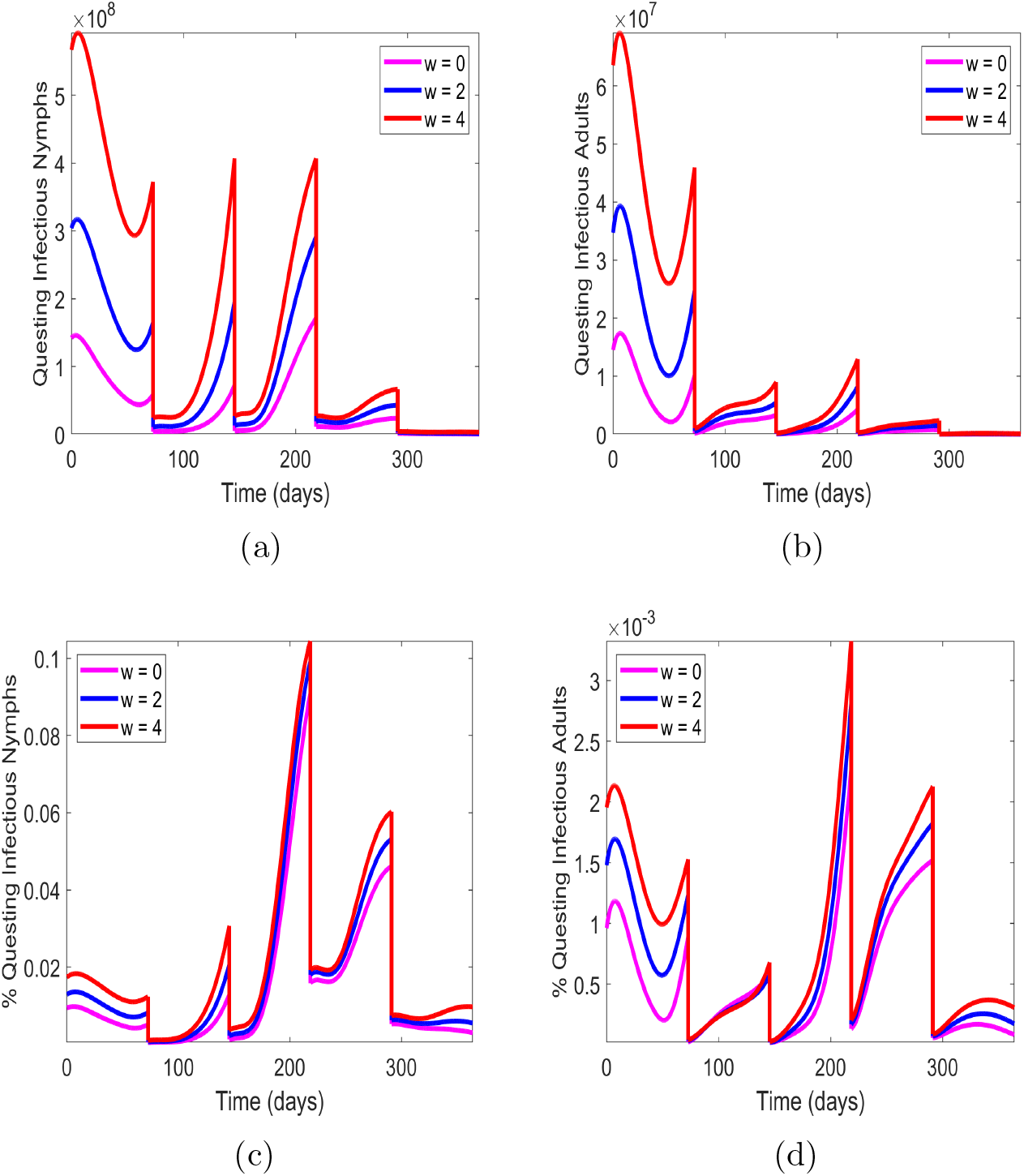
Nymph and adult tick population and disease dynamics under endemic scenario with four prescribed burns for current temperature *w* = 0 and increased temperature *w* = 2, 4 °C. (a) and (b) Questing infectious nymphs and adults; (c) and (d) Prevalence of questing nymphs and adults.

Because of the increased threat that ehrlichiosis poses, we wanted to investigate whether or not prescribed fire might be a viable solution to mitigating the increased prevalence of both ticks and disease. To evaluate this, we implemented four prescribed burns implemented evenly throughout the year. This is typically unnecessary in an environment that is typically managed using prescribed burns, but it may be done if the goal is to use prescribed burns as a means of tick population control. In this case, we see differences in how each tick-life stage responds following a burn depending on the time of year. We observed in in Figure 7(a), that the infectious questing nymphs have a similar profile of increase following the first and second burns and then very little increase as we approach the end of the year when less ideal temperatures occur. The nymph populations are reduced by two orders of magnitude by day 300 compared to Figure 6(a). We do still see an increased number of ticks as temperature increases, though the difference between each temperature increase is not as severe as the endemic case without fire and this difference diminishes as we reach the end of the year when colder temperatures occur.

Infectious questing adult ticks do not recover as readily as infectious questing nymphs following a prescribed burn. In Figure 7(b), we see an increase in the number of adults following the second burn, but even at the end of this increase, just before the next burn is implemented, the number of adults is roughly an order of magnitude less than in Figure 6(b) at around 200 days. We also see in Figure 7(b) that there is even less of a difference between the results for each increase in temperature compared to the nymphs in Figure 7(a). In Figure 7(c), we see that the percentage of questing infectious adults changes dramatically following each burn after an initial decrease. Following the first burn, the prevalence of disease slowly increases and there is almost no difference in prevalence as temperature increases. Following the second burn, there is a sharp increase in prevalence with a slight distinction as temperature increases. Following the third burn, the prevalence increases similarly regardless of temperature until day 250, after which, we see a greater increase in prevalence of disease as temperature increases. Following the final burn, there we see that the prevalence of disease increases as temperature increases, but from about day 330, it begins to decrease moving into the next year. We observe similar dynamics in Figure 7(d) for prevalence of adult ticks. These results indicate that prescribed fire remains an effective control method for nymph and adult ticks even with temperature increases of up to 4 °C and it also has the potential to reduce the prevalence of disease in ticks in an endemic situation.

#### 3.2.2 Invasion dynamics

Invasion of ticks into a new area is a situation that can occur when a female adult tick has finished feeding on a host that has traveled a significant distance while the tick was attached to a new area with little to no ticks. Examples of tick hosts that can travel significant distances before a tick has finished feeding are deer and migratory birds [35, 36, 37]. After the female has completed her blood meal, she detaches from the host and lays several thousand eggs. This can lead to new populations of ticks becoming endemic in new areas as well as the possibility of increased spread of tick-borne diseases since the ticks and/or their hosts may be infected [36, 37]. To explore these dynamics in the context of rising temperature, we present a simulation with disease and with only an initial number of eggs and a small number of feeding susceptible and infectious ticks and run it to determine the implications that invasion can have for the area.

From Figures 8(a) and 8(b), we see that the number of infectious questing nymphs and adults increases dramatically as temperature increases. They remain much lower in number than the endemic scenario, which indicates that ticks can take several years for these ticks to become fully established after being introduced to a new area. There is a five-fold increase in the peak number of infectious questing nymphs and adults, meaning that ticks will become more dangerous more quickly as temperature increases. The prevalence of disease in these ticks does not follow the same trend. We see in Figures 8(c) and 8(d) that the prevalence of disease reaches similar, though slightly lower levels compared to Figure 6(c) and Figure 6(d) within the first year. This is partially due to the fact that we assume the same level of prevalence in tick hosts as the endemic scenario. Since we are in an invasion scenario and ticks are only able to enter a new area via a host moving a significant distance, we assume those hosts have some level of infection a priori. One thing to note about this case is that the peak prevalence of disease is slightly lower as temperature increases. Since every tick population is growing significantly during this first year, and even faster as temperature increases, the rates of infection lag behind slightly and we don’t see them flip until roughly day 200 in the simulation. We do see that the prevalence of disease peaks faster and earlier on in the year as temperature increases, which is consistent with previous results [26]. This is especially dangerous in an invasion scenario where individuals are likely not aware of the ticks now present in the area and may not be checking themselves for ticks. The lower numbers of ticks in this scenario mitigate that danger temporarily, but ticks will become more and more prevalent each year, increasing the risk that they pose.

**Figure 8:**
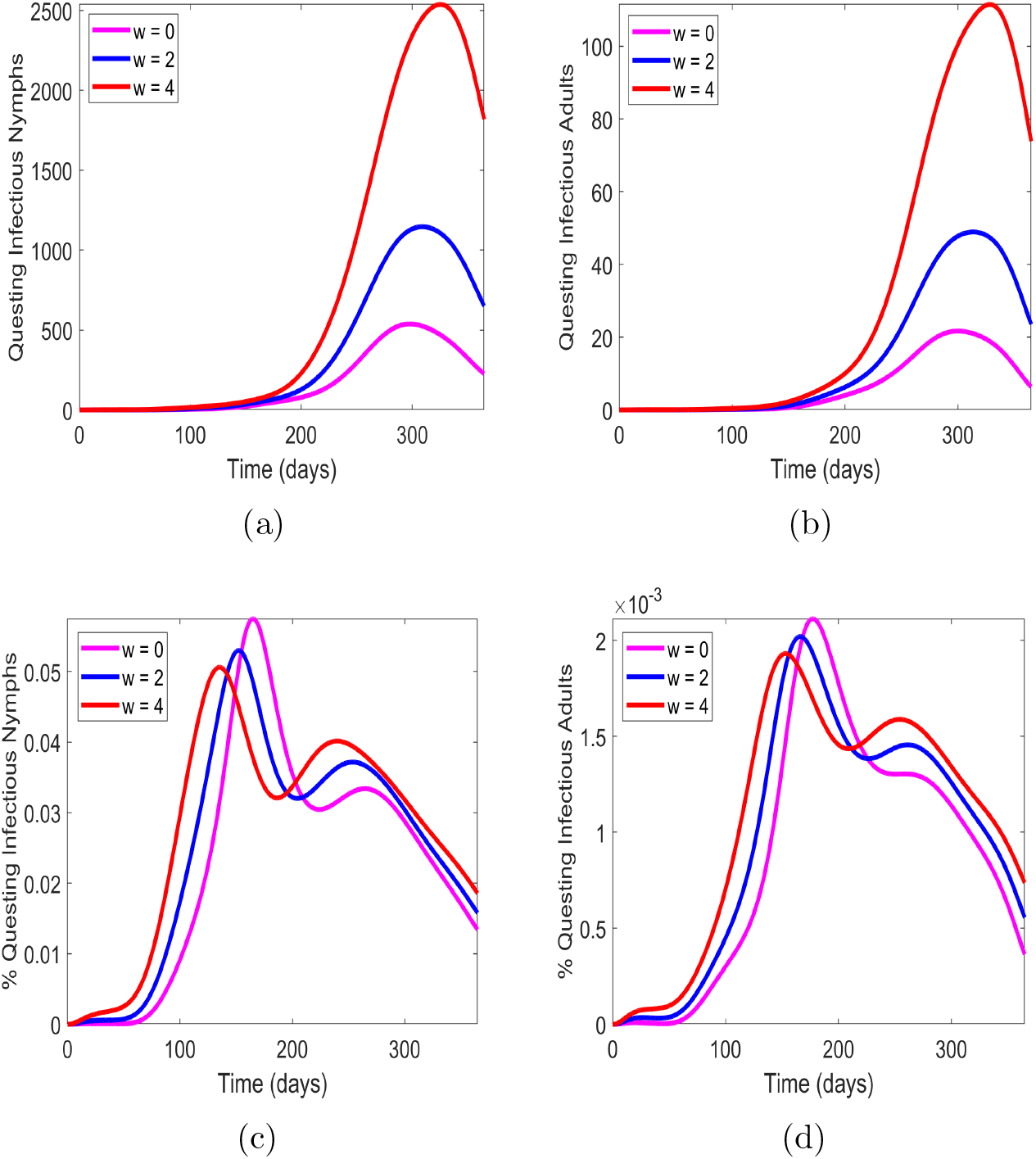
Nymph and adult tick population and disease dynamics in an invasion scenario for current temperature *w* = 0 and increased temperature *w* = 2, 4 °C. (a) and (b) Questing infectious nymphs and adults; (c) and (d) Prevalence of questing nymphs and adults.

For the following scenario in Figure 9, we present the results of implementing four prescribed burns in the area being invaded to explore the effect that burning has on an invasion and to determine if burning can be used to reduce the prevalence of disease in invading ticks.

**Figure 9:**
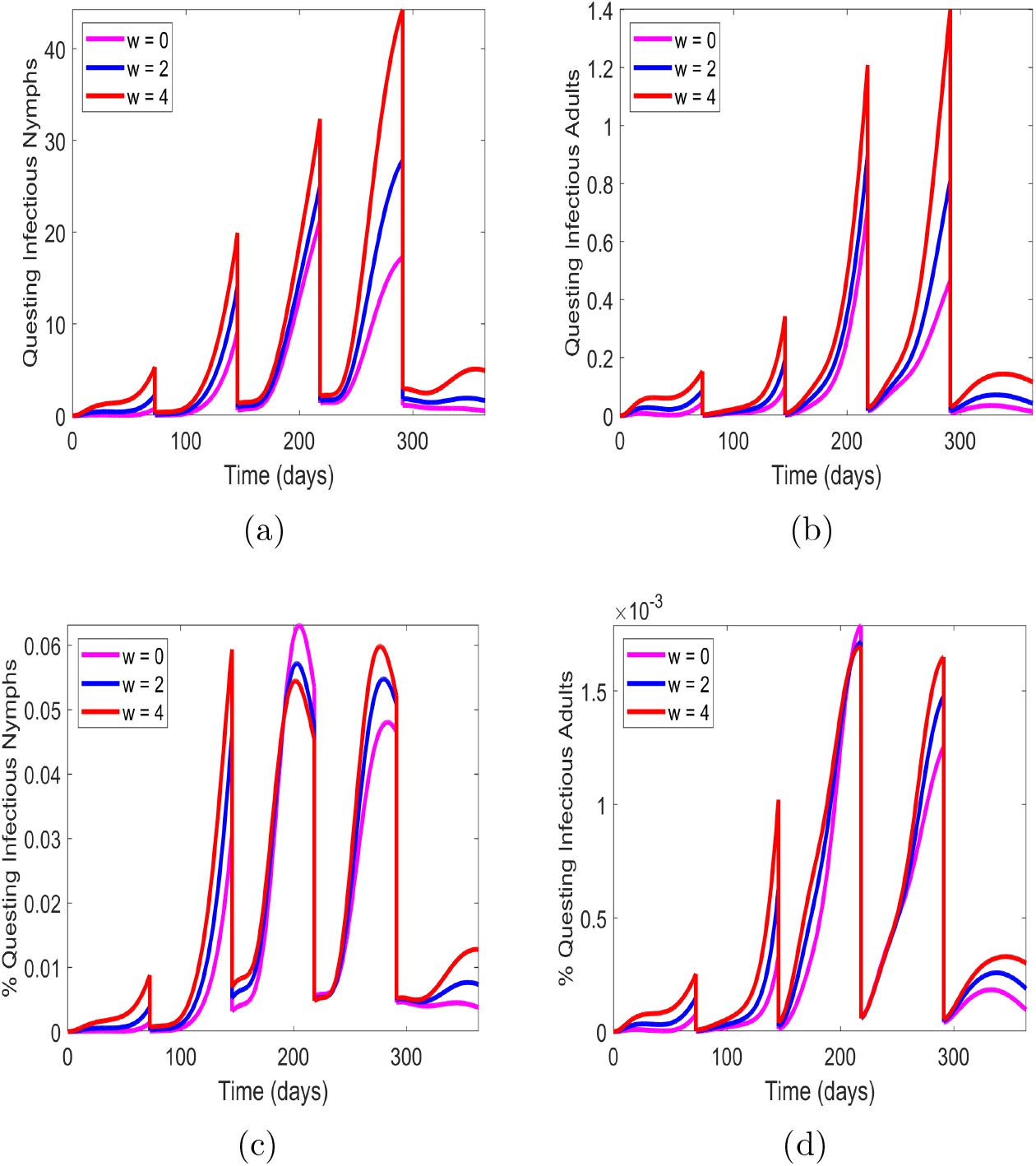
Nymph and adult tick population and disease dynamics in an invasion scenario with four prescribed burns for current temperature *w* = 0 and increased temperature *w* = 2, 4 °C. (a) and (b) Questing infectious nymphs and adults; (c) and (d) Prevalence of questing nymphs and adults.

Results indicate that prescribed burning is highly effective at slowing establishment of ticks (see Figure 9). Compared to Figure 8, the number of infectious questing nymphs has been reduced from *∼*2500 to *∼*50 at their peak in the case of *w* = 4 (see Figure 9(a)). Increases in the number of nymphs occur faster as temperature increases, but remain relatively low even at the largest temperature increase. The infectious questing adults have been reduced to near zero (Figure 9(b)) even at an increase of temperature of 4 °C, though this does not mean that they pose no threat as the survival of even a single female adult tick means that thousands of eggs may be laid to perpetuate the invasion of these ticks. In addition, we see that the prevalence of disease in both the nymph and adult ticks is again reduced immediately following a burn. From Figure 8(c), the prevalence of ehrlichiosis never surpasses *∼*0.06% for questing nymphs infected, whereas when prescribed burning is implemented, we see in Figure 9(c) that the infection prevalence surpass 0.06%, though it is important to keep in mind the low numbers of ticks present in this scenario; even if the prevalence of disease is slightly higher, the likelihood of a human encountering an infectious tick is drastically lower since almost none remain in the burned area. An interesting difference in this case is that the prevalence of disease in nymphs and adults is higher for *w* = 4 in most cases, except following the second burn (see Figures 9(c) and 9(d)). In other cases, the trends are similar to the endemic case with fire (see Figures 7(c) and 7(d)).

#### 3.2.3 Probabilistic implementation of prescribed fire

To provide a more realistic implementation of the initial effect that prescribed burning has on tick populations, we update *ν*_*i*_, *i* = *e, l, n, a* values before each burn is performed and set the value to a random value between the baseline and 30% below the baseline value (or above, as with the eggs). The variation in the effectiveness of prescribed burning can occur due to several reasons. Some of which include the escape of ticks on mobile hosts or deep into leaf litter that is not fully burned due to dampness. For a detailed description on how we chose the values of each *ν*_*i*_, *i* = *e, l, n, a* value for each burn, see Appendix A.

Figure 10 shows the results of a single simulation when the *ν*_*i*_, *i* = *e, l, n, a* parameters are determined in a probabilistic manner. From this, we see that following a prescribed burn, there is significant variation in how the number of infectious questing nymphs and adults recover following a burn compared to Figures 7(a) and 7(b). There is also significant variation in how the prevalence of disease changes following a prescribed burn compared to Figures 7(c) and 7(d). From Figures 10(a) and 10(b), we see that the trend of a higher number of ticks as temperature increases generally holds, except following the first and third burns in the questing infectious adult population for *w* = 4 and *w* = 2 temperatures, respectively.

**Figure 10:**
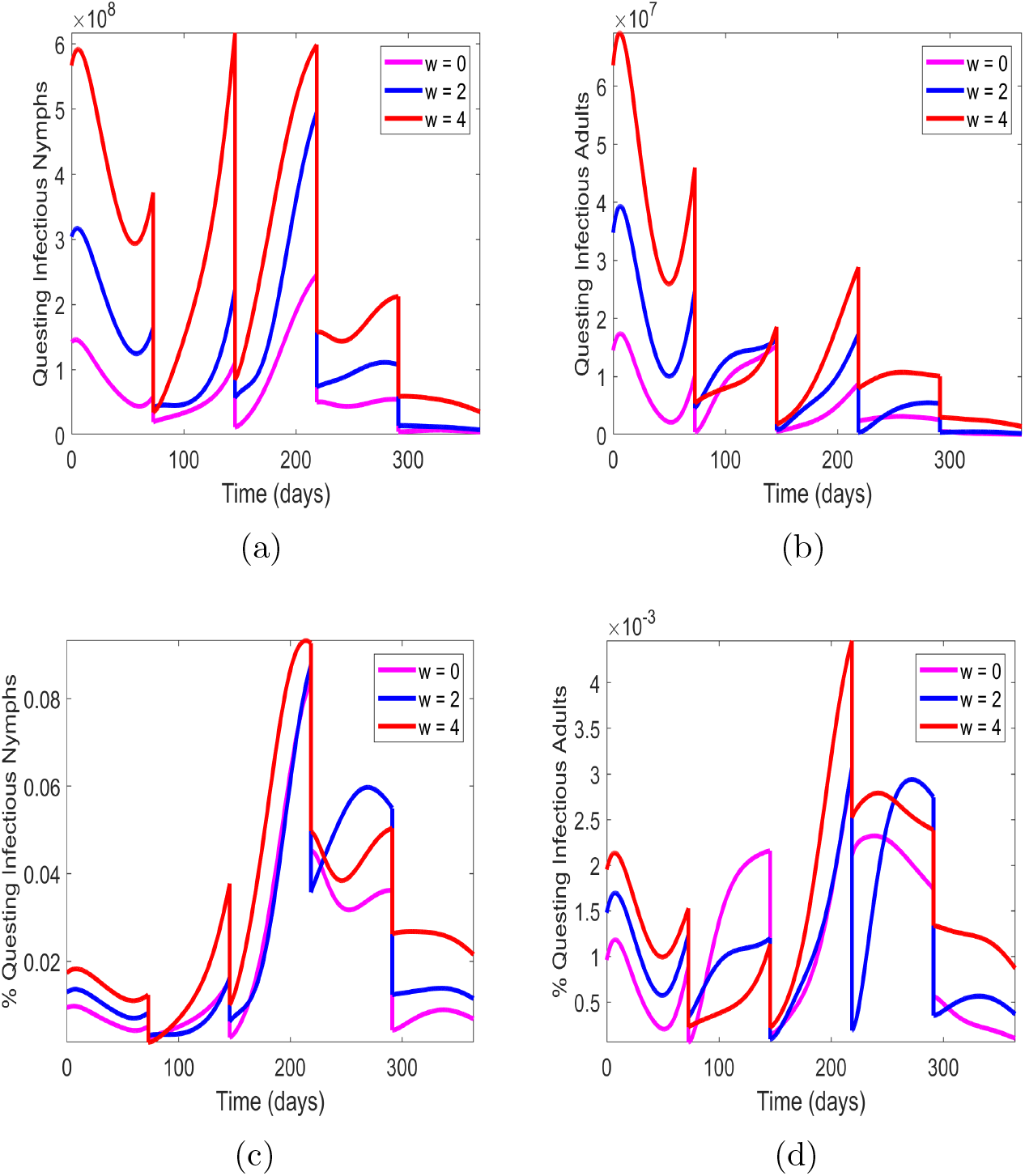
The effect of probabilistic prescribed fire under endemic scenario with four prescribed burns for current temperature *w* = 0 and increased temperature *w* = 2, 4 °C. (a) and (b) Questing infectious nymphs and adults; (c) and (d) Prevalence of questing nymphs and adults.

We also determined if these trends persist through multiple runs of the same scenario. Figure 11 below shows the results of the average of several simulations.

**Figure 11:**
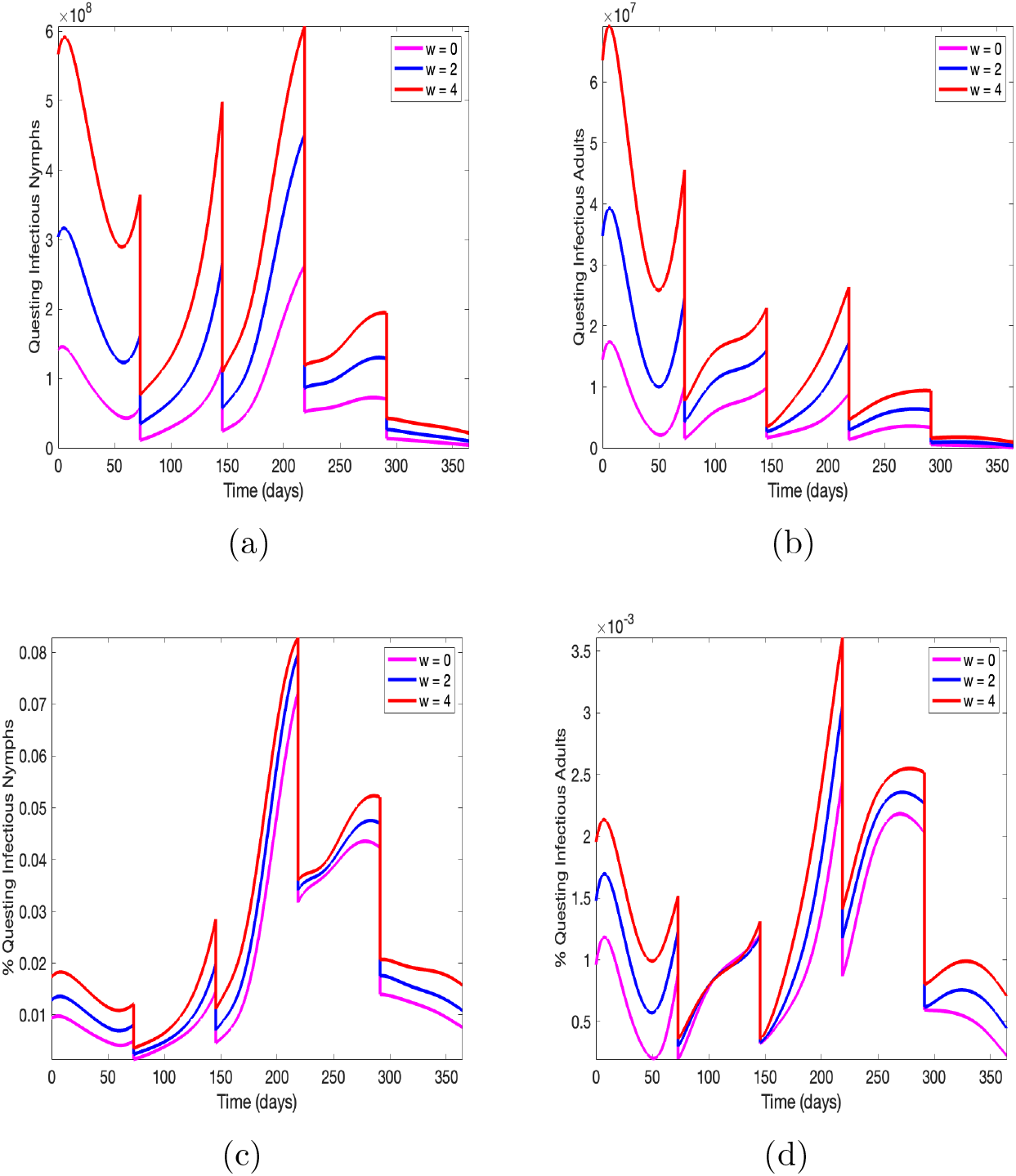
The effect of probabilistic prescribed fire under endemic scenario averaged over 100 run with four prescribed burns for current temperature *w* = 0 and increased temperature *w* = 2, 4 °C. (a) and (b) Questing infectious nymphs and adults; (c) and (d) Prevalence of questing nymphs and adults.

We ran 100 simulations using this probabilistic implementation of prescribed fire and subsequently averaged the results to see if the trends observed in Figure 10 held generally. The results of each run are displayed in Figure A1 in appendix A. From Figures 11(a) and 11(b), we see that nymphs and adults respond very differently following a prescribed burn depending on the time of year. Early on in the year (i.e. before 220 days), we see the infectious questing nymph population increase above pre-burn levels following the first and second burns. After the third burn, there is a relatively small increase in the number of infectious questing nymphs and once the final burn occurs, we see the population decrease until the end of the year. Infectious questing adult ticks remain in much larger numbers compared to Figure 7(b), and even exceed pre-burn levels following the second burn. The third burn leads to slight increases in each temperature case and there is little difference between the number of tick as temperature increases following the final burn. There are small, but clear increases in the prevalence of questing infectious nymphs in Figure 11(c) as temperature increases and these differences persist even immediately following a burn, unlike what we saw in Figure 7(c). Note that following each burn except the final, the prevalence of infectious questing nymphs increases consistently until the next burn. In Figure 11(d), we observed that the prevalence of infectious questing adult ticks remains surprisingly similar to the case with a static implementation of prescribed fire (see Figure 7(d)). Following the second burn, there is very little difference in the prevalence regardless of an increase in temperature, but this trend does not persist following the second and third burns. Following the final burn, we again see an increase in the prevalence until around day 330, after which the prevalence slowly decreases.

Figure 12 displays the results of a single simulation under an invasion scenario with probabilistic prescribed fire. From Figures 12(a) and 12(b), we see that prescribed fire is slightly less effective compared to the static case. We still observe that the higher the temperature, the more infectious questing nymphs and adults, but still at very low numbers compared to the invasion scenario without prescribed fire (Figure 8). The prevalence of infectious questing nymphs and adults shown in Figures 12(c) and 12(d) show significant variation. The prevalence of infectious questing nymphs is similar to the static case following the first burn, but following the second burn we see that an increase in temperature does not imply an increase in the prevalence of questing infectious nymphs as was seen in Figure 8(c) and this is also the case following the fourth burn as well. Interestingly, the prevalence of questing infectious adult ticks also responds similarly following the first burn, in that it increase consistently and there is a clear increase when going from *w* = 0, 2, and 4 °C. Following the second burn, we see that with current temperatures (*w* = 0) and temperature increase of 2 °C, a higher prevalence of infectious questing adults is achieved, but this is not the case following the third burn. The third burn leaves the base case (*w* = 0) at a higher initial prevalence, but this is quickly overtaken by the *w* = 2 and *w* = 4 °C cases, with the highest prevalence being achieved when *w* = 4 °C. After the final burn, the prevalence decreases for *w* = 0, and increases significantly for *w* = 2 before decreasing. For the highest temperature case (*w* = 4), there is a significant decrease.

**Figure 12:**
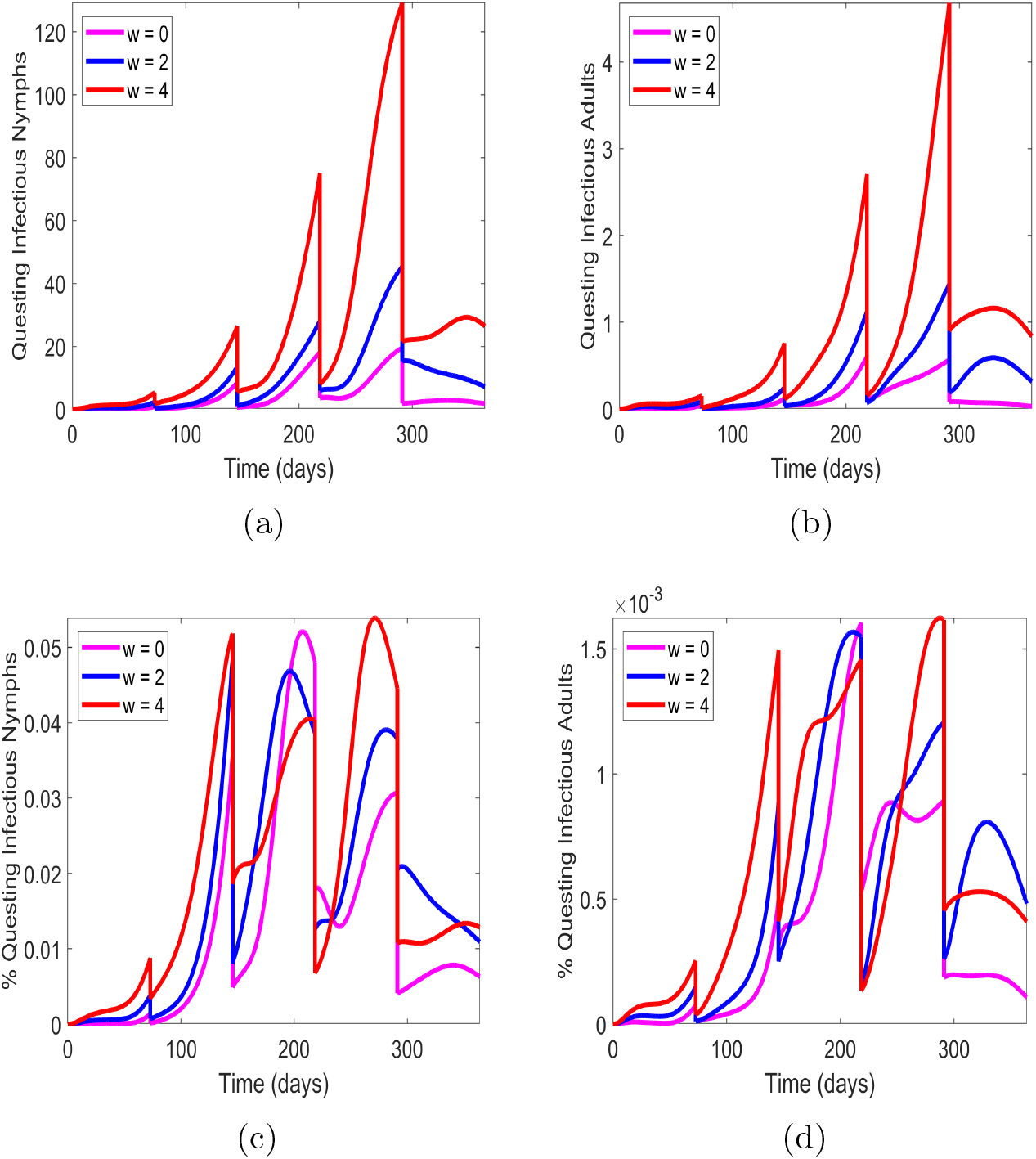
The effect of probabilistic prescribed fire in an invasion scenario with four prescribed burns for current temperature *w* = 0 and increased temperature *w* = 2, 4 °C. (a) and (b) Questing infectious nymphs and adults; (c) and (d) Prevalence of questing nymphs and adults.

The results of four probabilistic prescribed burns averaged over 100 runs in an invasion scenario is displayed in Figure 13. The results of each individual run are shown together in Figure A2 in Appendix A. As occurred in the endemic case, we see that much of the variation that was seen in a single run of probabilistic prescribed fire is not maintained when averaging over several runs. We again see in Figures 13(a) and 13(b) slightly higher numbers of infectious questing nymphs and adults compared to the static case, though it is still clear that invasion has been delayed thanks to the prescribed burns. In Figures 13(c) and 13(d) we observed that the prevalence of infectious questing nymphs and adults again have commonalities following the first burn in that they increase and there is a greater prevalence of infectious questing nymphs and adults as temperature increases. Following the second burn, there is much less of a difference in prevalence as temperature increases, though the peak reach for the prevalence of questing infectious nymphs is slightly higher for the *w* = 0 case, which was also observed in Figure 9(c). After the third and fourth burns, we see that the trend of increasing prevalence of infectious questing nymphs and adults as temperature increases holds true.

**Figure 13:**
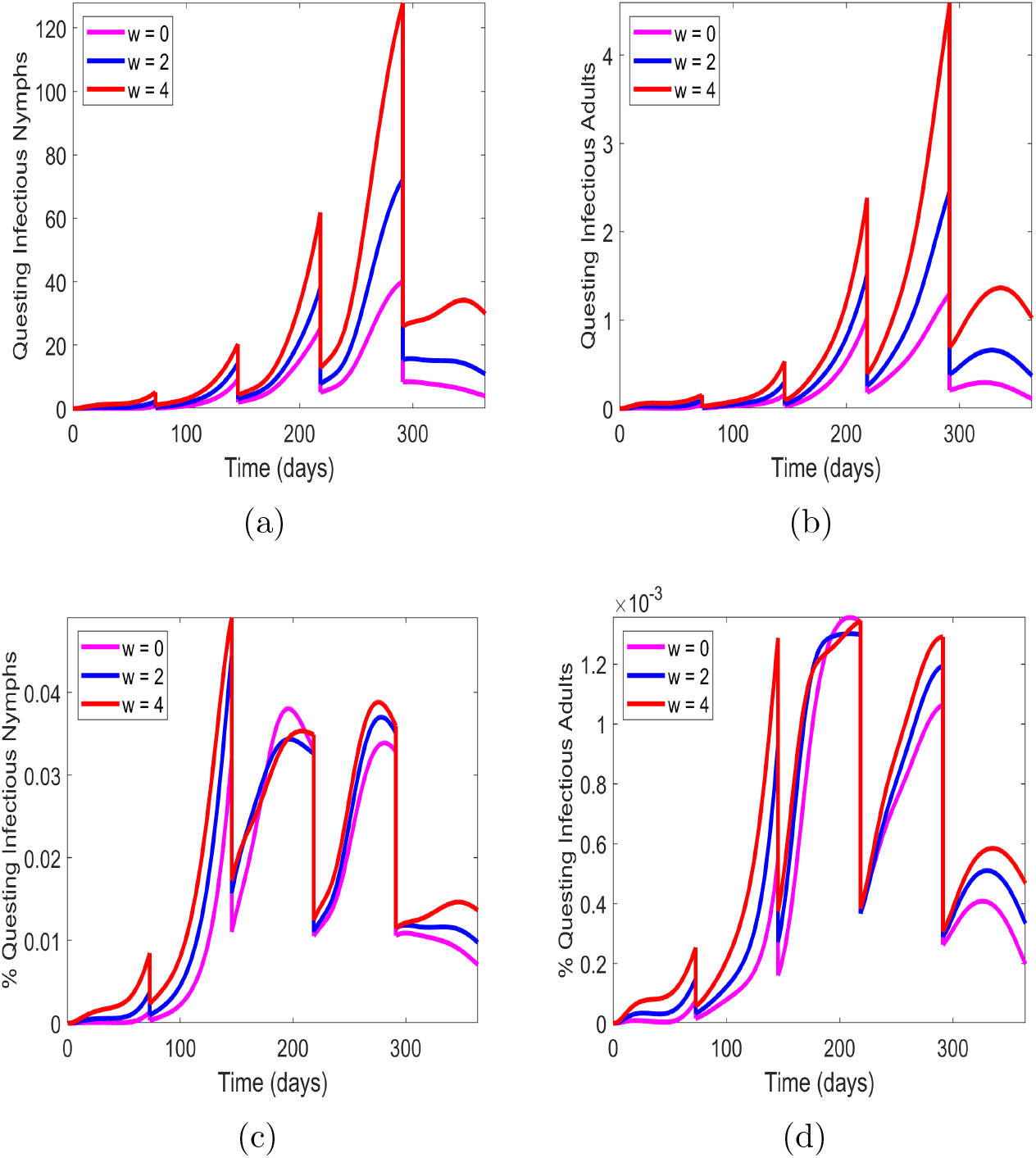
The effect of probabilistic prescribed fire in an invasion scenario averaged over 100 runs urrent temperature *w* = 0 and increased temperature *w* = 2, 4 °C. (a) and (b) Questing infectious nymphs and adults; (c) and (d) Prevalence of questing nymphs and adults.

## 4 Discussion

As far as we know, there have been two approaches used thus far to model tick populations and the effects of prescribed burning on those ticks [24, 25]. The first by Guo and Agusto [24] evaluated the effectiveness of high- and low-intensity prescribed burning as the duration between burns increases via an impulsive system of ordinary differential equations. They found that there was little difference in the number of *I. scapularis* ticks calculated in their simulations with a high- and low-intensity burn interval of six years (see Figure 4 in [24]). They also concluded that burn intensity matters more than frequency, which is important since high-intensity burns are likely not able to be performed as often as low intensity burns and thus one may opt for performing a high-intensity burn every few years rather than low-intensity burns more often. The second by Fulk et. al. [25] extended the model from [24] to a system of partial differential equations in order to determine how the effects of a spatial setting alter the effectiveness of prescribed burns. There, we found that a crucial determinant of the effectiveness of a prescribed burn was the size of the burn being performed. Since ticks and the hosts that they attach to tend to disperse across an environment, if the size of the burn being performed is smaller than the rate of movement of the ticks/hosts, the effectiveness of the burn may be diminished as they repopulate the burned area by diffusing from the surrounding area. Those models focused on *Ixodes scapularis*, the main vector for Lyme disease in much of the United States.

While it is likely that some of the conclusions hold for *Amblyomma americanum* and other tick species (e.g. high intensity burns being more effective than low intensity burns and smaller prescribed fires being less effective at reducing tick populations) since the ranges of some of these ticks overlap, the physiological differences between *Ixodes scapularis, Amblyomma americanum* and other tick species that are prominent in the United States (e.g. *Dermacentor variabilis*) warrant a thorough investigation into the differing effects of prescribed burning on each of these ticks. Gleim et. al. [22] provided support to this idea when she found that the distribution of various tick species in southwestern Georgia and northeastern Florida differed by site (e.g. unburned sites surrounded by unburned were dominated by *Amblyomma americanum*, while unburned sites surround by commonly burned areas were dominated by *Ixodes scapularis*).

Due to these findings, it thus important to include more aspects of this tick’s biology to properly determine the effect of rising temperature on populations of *Amblyomma americanum* ticks as well as the effects of prescribed burning in both endemic and invasion scenarios. Our results indicated that, depending on the life stage of the tick, populations of *Amblyomma americanum* may be able to recover very quickly following a prescribed burn in an endemic scenario, especially if the burn occurs before the ticks reach their highest peak numbers (see the increases in the nymph population in Figure 7(a) following the first and second burns), but frequent burning over the course of a year significantly reduces the number of ticks. Adult *Amblyomma americanum* ticks increased at lower rates following a burn (see Figure 7(b)), indicating that timing a prescribed burn when this life stage is most prevalent may be lead to increased effectiveness in reducing the number of ticks, especially in the following year since egg-laying adults are the main source of questing larvae.

The results of the invasion scenarios seem to indicate that as temperature increases, *Amblyomma americanum* will be able to establish itself in large numbers much more quickly. When prescribed fire is introduced into an area where these ticks have recently invaded, we see their populations grow at much lower rates. We also see that few infectious questing adult ticks are able to survive the burns, further indicating that timing a burn during times of the year when this life stage is most active may be effective at further slowing the establishment of these ticks. Eradication of ticks from an area using prescribed burning alone likely is not possible [25], thus additional control measures should be implemented in conjunction with prescribed burning if the goal is eradication of ticks. Four prescribed burns is an unsustainable number to perform on an annual basis, but it is extremely effective for an initial reduction in tick populations in both endemic and invasion scenarios. If the goal of prescribed burns is tick population management, then as many burns as possible should be performed in a year as possible, followed by additional an burn at regular intervals (annually, or more frequently if possible).

The prevalence of infectious questing nymphs and adults was also evaluated in each of our scenarios. The percentage of infectious questing nymphs and adults increases slightly in an endemic scenario as temperature increases, but the greater threat is the increase in the number of infectious questing nymphs and adults (see Figure 6). The invasion scenario highlighted another likely effect of rising temperatures. Prescribed burning seems to be initially effective at reducing the prevalence of infectious questing nymphs and adults, however the prevalence quickly returns to and exceed pre-burn levels if the burn is performed before approximately day 220 for nymphs. The prevalence of infectious questing adults does not increase in the same way that the nymphs do, meaning that an individual is less likely to encounter an infectious questing adult in both endemic and invasion scenarios and with or without prescribed burning. These factors indicate that prescribed burning may not be the most effective way to reduce the prevalence of disease in tick populations. This is because prescribed burning likely does not kill many of the reservoir hosts of ehrlichiosis. One of the key reservoir hosts is white-tailed deer, which are able to move quickly relative to a prescribed burn and may return to recently burned areas quickly to feed on newly growing vegetation. If eradication of disease is the main goal in an area, then other control methods that directly target these reservoir hosts may lead to a more effective long-term reduction in the prevalence of disease in *Amblyomma americanum*.

## 5 Conclusion

In this study we investigated the effect of rising temperature on *Amblyomma americanum* populations and the prevalence of infectious questing nymphs and adults under endemic and invasion scenarios. We also evaluated the effectiveness of prescribed burning tick populations and the percentage of infectious questing nymphs and adults. We have three main conclusions on the effects of rising temperature in the absence of prescribed burning: (1) as temperature increases, there are significant increases in the number of infectious questing nymphs and adults. (2) The prevalence of infectious questing nymphs and adults increases slightly, but the greater threat to public health is the increase in the number of infectious ticks. (3) Ehrlichiosis becomes established in the questing nymph and adult tick populations quicker under the invasion scenario as temperature increases.

In the presence of prescribed burning, we found that infectious questing nymph populations increase sharply following a prescribed burn if the burn is performed earlier in the year (i.e. before 220 days) in an endemic scenario whereas infectious questing adults did not increase as significantly following a burn at any point during the year. Prescribed burning also had an initial effect on the prevalence of infectious questing nymphs and adults, but these quickly recovered to similar levels as the cases without prescribed burning, though the number of infectious questing ticks was dramatically reduced which is more important from a public health perspective since this decreases the encounter rate with humans. We also note that these conclusions with prescribed burning hold regardless of increases in temperature, indicating that prescribed burning remains an effective control method for *Amblyomma americanum* even in the case of a significant increase in temperature of 2 and 4 °C.

There are some limitations to our model that should be mentioned. We assumed a constant number of susceptible and infectious small and large hosts. We do not expect that dynamic populations of these hosts would lead to significantly different conclusions since one of the main hosts for *Amblyomma americanum* (and ehrlichiosis) in many areas is the white-tailed deer, which remains active to varying degrees during much of the year. We hypothesized that timing a prescribed burn to when the adult tick population is near its peak may lead to increased effectiveness of the burn in reducing tick populations in the following year. In Section 4, though we are not able to address that question directly with our current model since our goal was not to accurately model tick phenology. This is a very interesting question that likely has a different answer depending on which tick species is being considered and we plan to address it in future works.

## Data Availability

All data and code produced in this study are available upon reasonable request to the corresponding author

## A Probabilistic Prescribed Fire

To calculate the proportion of ticks reduced following each of the prescribed burns implemented in Section 3.2.3, we used the following scheme. For the egg population, we first defined the range of a uniform distribution on the interval [0, 0.3] for a maximum reduction of 30% and extracted a value from this range at each burn throughout the year. For the other tick populations, we used ranges for a uniform distribution from 30% below to the baseline values described in Section 2.3. So, for the larvae populations, the range of possible values for the uniform distribution was [0.693, 0.99]. For the nymph populations, the range of possible values for the uniform distribution was [0.653919, 0.93417]. For the adult populations, the range of possible values for the uniform distribution was [0.6846392, 0.978056]. These values are used in the impulsive equations described in Section 2.2.

**Figure A1:**
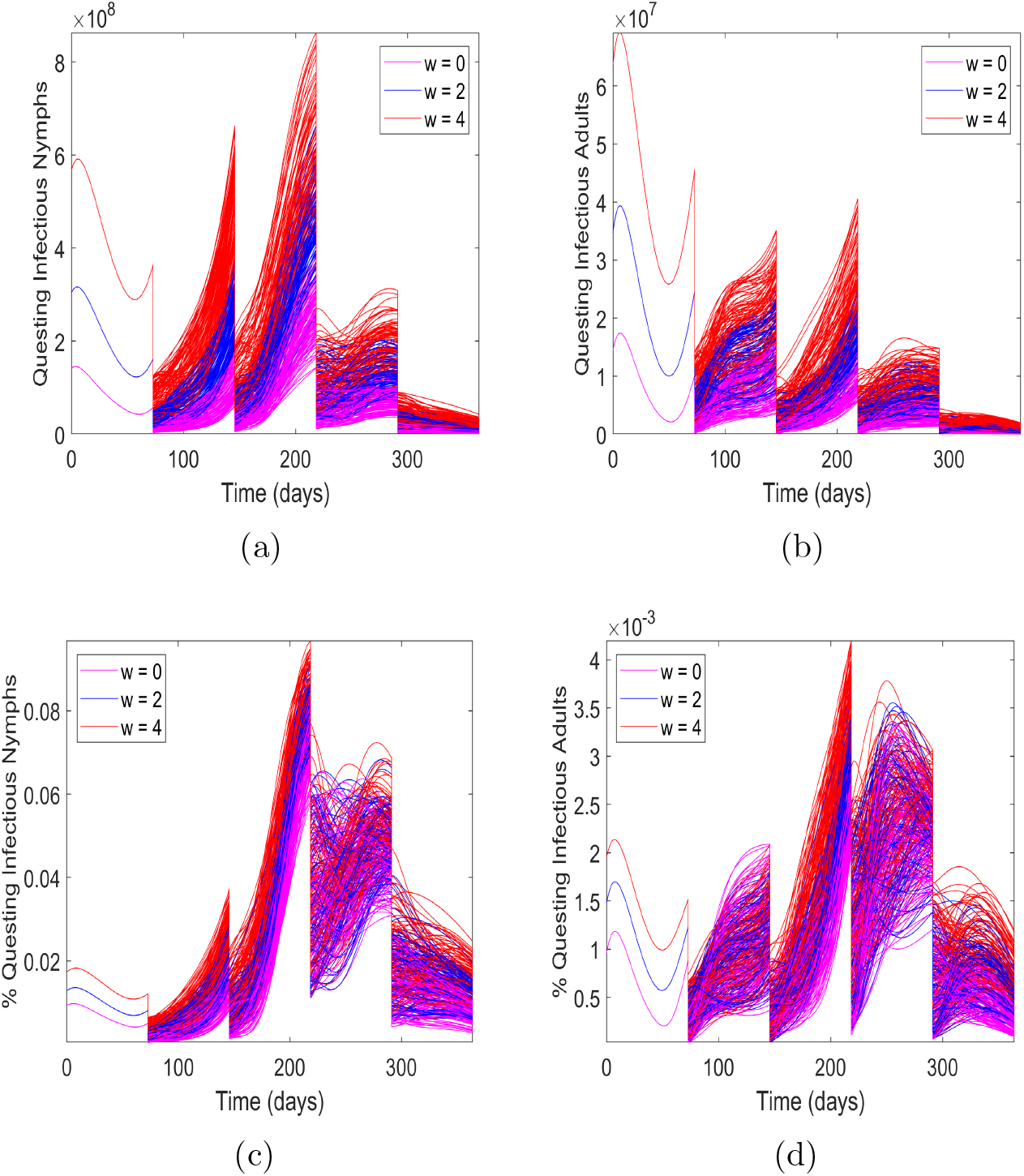
The effect of probabilistic prescribed fire in an endemic scenario over 100 runs with four prescribed burns for current temperature *w* = 0 and increased temperature *w* = 2, 4 °C. (a) and (b) Questing infectious nymphs and adults; (c) and (d) Prevalence of questing nymphs and adults.

Figure A1 shows the results of each run used to generate the average effect of probabilistic prescribed burning provided in Figure 11. Burns initiated earlier in the year lead to similar increases in tick nymphs as temperature increases. Following the third burn, there is more variation, particularly in the prevalence of disease in tick nymphs. There is much more variation seen in the numbers of adult ticks following the first burn, but not the second. This also holds for the prevalence of disease in the tick populations. Following the third and fourth burns, we again see significant variation. When averaging over all 100 runs, this variation is not maintained.

**Figure A2:**
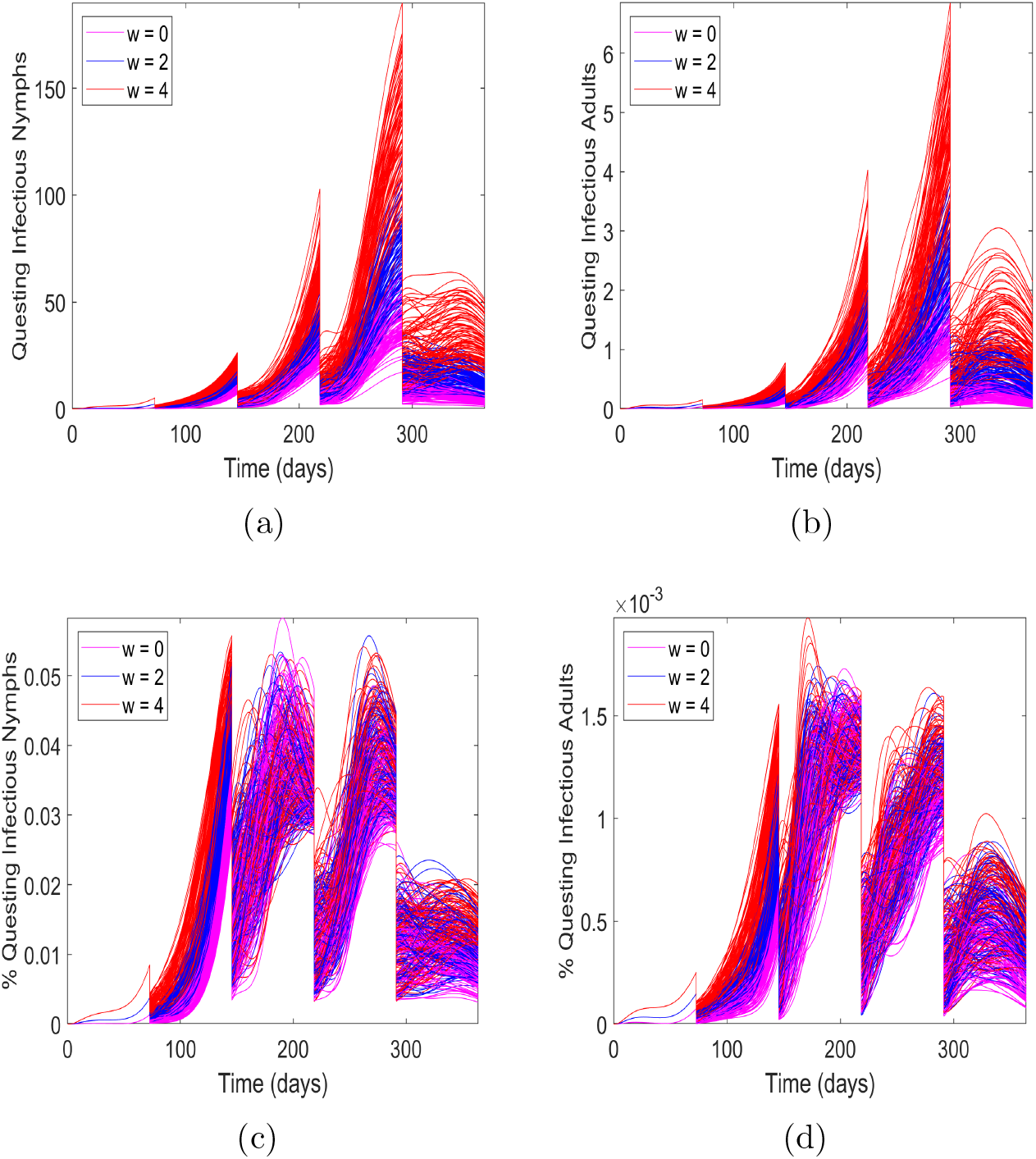
The effect of probabilistic prescribed fire in an invasion scenario over 100 runs with four prescribed burns for current temperature *w* = 0 and increased temperature *w* = 2, 4 °C. (a) and (b) Questing infectious nymphs and adults; (c) and (d) Prevalence of questing nymphs and adults.

Figure A2 shows the results of each run used to generate the average effect of probabilistic prescribed burning provided in Figure 13. There is a clear distinction between the numbers of infectious questing nymphs and adults as temperature increases throughout the year, which was not observed in the endemic scenario. Prevalence of infectious questing nymphs and adults varies significantly following the second third and fourth burns, but not following the first burn.

